# Barriers to and enablers of quality improvement in primary health care in low- and middle-income countries: a systematic review

**DOI:** 10.1101/2023.07.29.23293377

**Authors:** Camlus Odhus¹, Ruth Kapanga², Elizabeth Oele³

## Abstract

The quality of health care remains generally poor across primary health care settings, especially in low- and middle-income countries where tertiary care tends to take up much of the limited resources despite primary health care being the first (and often the only) point of contact with the health system for nearly 80 per cent of people in these countries. Evidence is needed on barriers and enablers of quality improvement initiatives. This systematic review sought to answer the question: What are the enablers of and barriers to quality improvement in primary health care in low- and middle-income countries? It adopted an integrative review approach with narrative evidence synthesis, which combined qualitative and mixed methods research studies systematically. Using a customized geographic search filter for LMICs developed by the Cochrane Collaboration, Scopus, Academic Search Ultimate, MEDLINE, CINAHL, PSYCHINFO, EMBASE, ProQuest Dissertations and Overton.io (a new database for LMIC literature) were searched in January and February 2023, as were selected websites and journals. 7,077 reports were retrieved. After removing duplicates, four reviewers screened titles, abstracts, and full texts, and performed quality appraisal and data extraction and synthesis. 50 reports from 47 studies were included, covering 52 LMIC settings. Six themes related to barriers and enablers of quality improvement were identified and organized using the model for understanding success in quality (MUSIQ) and the consolidated framework for implementation research (CFIR). These were: microsystem of quality improvement, intervention attributes, implementing organization and team, health systems support and capacity, external environment and structural factors, and execution. Decision makers, practitioners, funders, implementers, and other stakeholders can use the evidence from this systematic review to minimize barriers and amplify enablers to better the chances that quality improvement initiatives will be successful in resource-limited settings. PROSPERO registration: CRD42023395166.

## Introduction

This review synthesizes literature on barriers to and enablers of efforts by health workers and different stakeholders to improve the quality of primary health care in low- and middle-income countries.

The Agency for Healthcare Research and Quality (AHRQ) contends that “Quality improvement (QI) is essential to achieving the triple aim of improving the health of the population, enhancing patient experiences and outcomes, and reducing the per capita cost of care, and to improving provider experience” [1].

Improving the quality of primary health care is fundamental to the achievement of health goals in lower-middle-income countries [2]. Primary health care (PHC) is all too important because up to eight in every ten people in low- and middle-income countries (LMICs) depend on it to meet their health and care needs [3]. For the better part of modern healthcare history, the poor quality of healthcare has generated concerns among practitioners, researchers, and policymakers [4-7] and those concerns have only grown louder [2]. This is because despite more, though increasingly uncertain, investments and rapid innovation, health outcomes have stagnated with rising inequalities in many LMICs [8, 9] potentially leading to wastage, harm, and even preventable deaths [10].

While barriers (constraints or limitations) prevent the realization of full benefits from quality improvement (QI) interventions, enablers (also known as promoters, facilitators, or motivators) unlock the potential of such interventions and typically enhance the desired level of quality of primary health care. Both range from the individual or micro (e.g., nurse manager knowledge and behaviour), to institutional-organizational or meso (e.g., shared beliefs, attitudes and practices at a health centre or hospital), to system-wide and societal or macro influences, e.g., implicit, or explicit values that drive QI culture, priority-setting, or investments.

First, it is necessary to define key terms. Primary health care (PHC) is challenging to define because it includes or precludes different packages of health services in different contexts. Perhaps it is due to this challenge that the World Bank, the World Health Organization and others [3] opted to define PHC rather broadly as “a health- and social-service delivery platform or system uniquely designed to meet communities’ health and health care needs across a comprehensive spectrum of services— including health services from promotive to palliative—in a continuous, integrated, and people-centred manner.” PHC services are often attuned to the prevailing socioeconomic, political and historical contexts of communities, in addition to the financial and health workforce considerations in the given country setting [11, 12].

Competing but comparable definitions of quality of care which hold important implications for how quality improvement (in healthcare) is defined and operationalized have been proposed by the World Health Organization [13], by the United States National Academy of Medicine, formerly Institute of Medicine or IOM [14], and others [15, 16]. However, consensus remains elusive [15]. But, QI - with roots in manufacturing in 1920s – can be defined as a framework with tools, approaches, techniques, and skillsets including assessment and measurement, goal-setting, and shifts in mindsets geared towards improving equity, access, effectiveness, patient-centeredness, and safety of healthcare [16, 17]. Ongoing debates on the level (individual or population), scope (bounded setting or whole systems), and approaches (evidence-based practice, multidisciplinary) to healthcare quality improvement are unlikely to be concluded soon [18-20]. The review considered the lack of consensus by being as inclusive as possible, avoiding a one-size-fits-all approach to defining quality improvement.

Some of the existing reviews have synthesized evidence on patient safety culture in Latin American Hospitals [21], barriers and enablers to the provision of emergency obstetric care in Nigeria [22] and in LMICs [23], and interventions to improve anti-retroviral therapy programmes in sub-Saharan Africa [24]. A COCHRANE review studied the use of reminders in health care [25]. Notably, an umbrella review [26] describing the influence of contextual factors on hospital QI using the Model for Understanding Success in Quality (MUSIQ) tool [27] found that previous systematic reviews overwhelmingly included studies from high income countries in North America, Europe, and Southeast Asia and very few from LMICs (Egypt, South Africa, Zambia, Sudan, Costa Rica, Brazil, and Argentina). A more recent realist-inspired review [28] confined itself to a specific type of QI, namely “QI collaboratives” to investigate contexts, mechanisms and outcomes but still included only five (out of 32) primary studies from LMICs. Still, other reviews have confined themselves to ‘training and measurement’ [29-31] and patient safety education [32, 33]. No systematic review was found that synthesized literature from low- and middle-income countries to inform holistic quality improvement policy and practice specifically in primary health care.

### Review aim and questions

The systematic review aimed to describe the barriers to and enablers of quality improvement (QI) within primary health care in low- and middle-income countries (LMICs). The review sought to answer the following three closely related questions:

1. What are the barriers to and enablers of QI in primary health care in low- and middle-income countries?
2. What is the shared knowledge, beliefs, values, attitudes, and practices (collectively called ‘culture’) of LMICs’ health workers and stakeholders regarding QI in primary health care?
3. What micro (individual or personal), meso (institutional or organizational) and macro (societal or structural) factors motivate health workers and managers involved in primary health care QI in LMICs?

## Review approach and methods

### Review approach

An integrative approach [34] incorporating narrative synthesis [35] for this systematic literature review. Integrative reviews are suitable for combining studies from disparate methodological approaches such as mixed methods and qualitative studies explicitly and has played an expanding role in health systems and policy research [34], contributing to evidence-based policy and practice. The framework for integrative review commences with problem identification, proceeds through a literature search, appraisal of data and analysis, before concluding with data presentation. A narrative approach to evidence synthesis relies on ‘storytelling’, as its name suggests, and is commensurate with the overall integrative review approach [35]. Correctly performed, narrative synthesis can minimize bias in reviews, ensuring that the eventual review output can be trusted by policymakers and practitioners alike. In the present review, this approach was used to enrich the data analysis and presentation stages of the integrative review. The findings of this systematic review incorporating primary studies on different aspects of QI were synthesized narratively. To comprehensively answer the review question, both mixed methods and qualitative studies investigating barriers, enablers, culture, and other contextual influences on diverse quality improvement interventions in primary health care in low- and middle-income countries were included.

### Literature search strategy

The search for primary research reports was performed in January and February 2023. Electronic databases (MEDLINE, PSYCHINFO, EMBASE and CINHAL) were searched using a mix of free-text (key words) and Medical Subject Headings (MeSH terms), refined for each database using EBSCO interface. More search explored TRIP, Academic Search Ultimate, Web of Science, Scopus, and Africa Index Medicus. The key terms used to develop the literature search strategy drew upon the SPIDER mnemonic [37] included “Quality Improvement” AND “Primary Health Care” AND “Low-middle-income countries”.

A scoping search was first used to check how studies are indexed and the relevant key words and synonyms. It was also used to test and refine the search strategy. A priori search strategy was then developed and applied to each database flexibly. A sample search strategy used for MEDLINE is contained in supplementary file (S3 Fig). Neither time nor language filters were applied at this stage. Boolean and near field operators were used to expand and narrow the search as appropriate. A geographic search filter for LMICs developed by the Cochrane Collaboration’s Effective Practice and Organisation of Care (EPOC) group [36] was applied to exclude high income countries. Literature was searched and retrieved in January and February 2023.

Grey literature including dissertations and thesis reports were sought from PROQUEST and the WHO and UNICEF public websites were also searched as was the preprint server, Medrxiv. To further reduce publication bias, Overton.io (an open research initiative to expand access to grey literature from LMICs) was also searched for grey literature. Finally, selected QI-focused journals (Health Policy and Planning, Implementation Science, International Journal for Healthcare Quality, BMJ Open Quality, Journal for Healthcare Quality, BMJ Quality and Safety, Journal of Health Services Research) were hand-searched as were reference lists of systematic reviews in the field of QI.

### Study selection

All (n=7,077) reports were imported into Rayyan systematic review management (web platform) where (n=4,110) duplicates were removed automatically and manually. Titles and abstracts (n=2,967) were screened by two reviewers independently and included (n=227) if they were deemed relevant. Conflicts throughout the selection process were resolved by consensus and did not require the intervention of a third person. At full text review, reports were read multiple times and subjected to inclusion and exclusion criteria as shown in Table 1. Inclusion and exclusion criteria were derived from the SPIDER mnemonic [37] and signified the information power of the primary research report to contribute answers to the review question(s).

**Table 1.** Inclusion and exclusion criteria.

| SPIDER element | Include | Exclude |
| --- | --- | --- |
| Sample | Facility-based HCWs<br>Community-based health workers<br>Health managers, policymakers and stakeholders across the health system | Exclude if others included and lumped alongside these in findings. |
| Phenomenon of interest | Quality improvement (not just quality of care or general health systems capacity or situation assessment)<br>Quality includes safety, effectiveness, patient-centeredness, timeliness, efficiency, equity in health uptake/access, utilization, or outcomes.<br>Must be primary care or primary health care oriented, reported separately from tertiary and referral level. | Exclude high income country context, exclude health technology assessments, exclude other systematic and umbrella reviews. Also exclude very low-quality studies (judged by consensus) and those from tertiary care (university/teaching and research hospitals).<br>Exclude editorials and opinion pieces, economic evaluations, and clinical case reports.<br>Include only primary empirical research (mixed or qualitative) reporting enablers to and barriers of quality improvement from perspective of health workers, health managers or regulators as study participants. |
| Design | Mixed methods and Qualitative designs.<br>Mixed methods papers have qualitative data detailing enablers or barriers. | Quantitative design with no discernible data on contextual drivers of QI measured or reported |
| Evaluation | Intervention to improve quality of health care i.e., efforts introduced to Change quality from level X to Y or measured from time X to time Y i.e., a QI initiative rather than just a measurement of quality of care. | Economic evaluations with no accompanying contextual data<br>One-off measurement seeking to perceptions of stakeholders on quality of care rather than on QI intervention/initiative/ project |
| Research type | Qualitative data reported separate from quantitative findings in mixed methods.<br>Qualitative research findings qualitatively reported (not quantified in percentages or numerical values).<br>Semi-structured or in-depth Interviews, focus groups, observation, ethnography etc. | Surveys, Randomized Trials with no process evaluations reporting barriers or enablers of QI initiative or QI project |

Eventually, 50 research reports from 47 studies were found that met the inclusion criteria for the systematic review following independent decisions by both reviewers. Fig 1 is a PRISMA flow chart showing results of the study selection process [38].

**Fig 1.** PRISMA Flow Chart.

### Assessment of study quality and relevance

The Mixed Methods Appraisal Tool, MMAT, checklist [39] was used to critically assess the quality of all 50 included full text reports prior to data extraction. MMAT checklist was especially suitable because it was developed for systematic reviews incorporating primary studies from different designs (qualitative, quantitative, and mixed methods). The first two screening questions ask whether there were clear research questions and if the data collected allowed the primary researchers to address the study’s research question. For qualitative studies the tool has five themes (with yes, no, or can’t tell response options): coherence between methodology and research question, coherence between data collection methods and research question, adequacy of findings given the data, coherence between the interpretation of findings and the data, and coherence in the research cycle from data sources, collection, analysis, and interpretation. To assess mixed methods studies MMAT focuses more on the appropriateness of mixing methods, whether the various methodologies were suitably combined, and how rigour and trustworthiness for each research tradition was maintained in the primary research. Each quality criteria entails “yes”, “no”, or “can’t tell” response options. In keeping with best practice for integrative reviews and narrative synthesis, no scoring was done, and no study was excluded from the analysis based on the results of the critical appraisal, but the strengths and limitations of each study were considered in the ensuing synthesis.

### Data extraction

The lead author extracted data from all 50 included reports while a second reviewer independently extracted data from a sample of 23, about half of all included reports. A comparison of both datasets showed no major inconsistencies. The bespoke data extraction form also had sections to capture QI theory (of change), description of the QI intervention, study setting, sample and population, barriers, and enablers as well as motivations and other contextual influences. Lastly, data on study conclusions, limitations and strengths, and recommendations (where available) were included. Data extraction made use of Microsoft Office Forms, hosted online.

### Data analysis

Data analysis involved the use of two frameworks commonly applied in Quality Improvement (QI) research. The Model for Understanding Success in Quality (MUSIQ) developed by [27] was predominantly used, complemented with the Consolidated Framework for Implementation Research, CFIR, [40]. MUSIQ contains concepts for understanding the external context of QI interventions while CFIR complemented this by providing a way to organise attributes intrinsic to the QI intervention itself. Concepts from these two frameworks were deductively applied to the entire dataset of 50 research articles and inductive coding with labels grounded in the data was done where data did not fit into the a priori coding framework. The entire process was iterative with multiple revisions. Atlas.ti version 9 (ATLAS.ti Scientific Software Development GmbH, Berlin) was used for coding and categorization.

First, to enable detailed and systematic analysis of this large dataset and in concert with the integrative review approach, studies were classified and grouped by geographic region, country income status and study topical focus. This allowed systematic comparison of studies and integration of their findings. Next, deductive codes from MUSIQ and CFIR were applied to the data extracted from studies in addition to new (inductive) codes. Codes were then grouped into categories (still informed by MUSIQ and CFIR) before being displayed in tables and matrices and network diagrams. Through comparisons and contrasts, noting surprising or unique findings and variability within and across subgroups, the analysis moved into the final phase. Here, a description of patterns in the form of themes concluded the analysis by narratively synthesizing subgroup patterns into an overall picture to address the review’s three aims: to describe the evidence on barriers and enablers of primary healthcare quality improvement; to uncover individual motivations (of health workers, managers and other stakeholders) for undertaking QI; and to describe what the culture (shared knowledge, language, or artifacts) of QI looks like in PHC settings in LMICs contexts. Verification of results was done by going back to primary studies to ascertain the link with eventual conclusions (S2 Table).

## Results

### Characteristics of included studies

Fifty primary research reports were included in the analysis. Twenty-eight had mixed methods design while 22 were qualitative, as summarized in Table 2. Signifying increasing interest in primary health care quality improvement by researchers, 41 of the studies were published in the last five years (2018 to date) while only nine were reported between 2012 and 2017. Although the review had aimed to include studies since 2000, none of the included studies covered the period 2000 to 2011. Turning to geographical coverage (S3 Table) for this systematic review that sought literature from low- and middle-income countries (LMICs) on barriers and enablers of quality improvement in relation to primary health care, fourty-one of the studies were based in sub-Saharan Africa (SSA), seven from Asia and two from Latin America (Costa Rica which is upper middle-income and Haiti which is lower middle-income). All seven studies based in Asian countries came from lower middle-income settings (India had three while Indonesia, Tajikistan, Papua New Guinea, and Sri Lanka had one study each). Out of the fourty-one studies from SSA, 37 reported research conducted in a single-country set-up while four covered multiple countries. In total, research reports covered 45 SSA countries. A close examination revealed that three publications [41, 42] were likely from the same QI intervention in Tanzania and Uganda and a further two publications [43, 44] were from the same project in Nigeria.

**Table 2.**
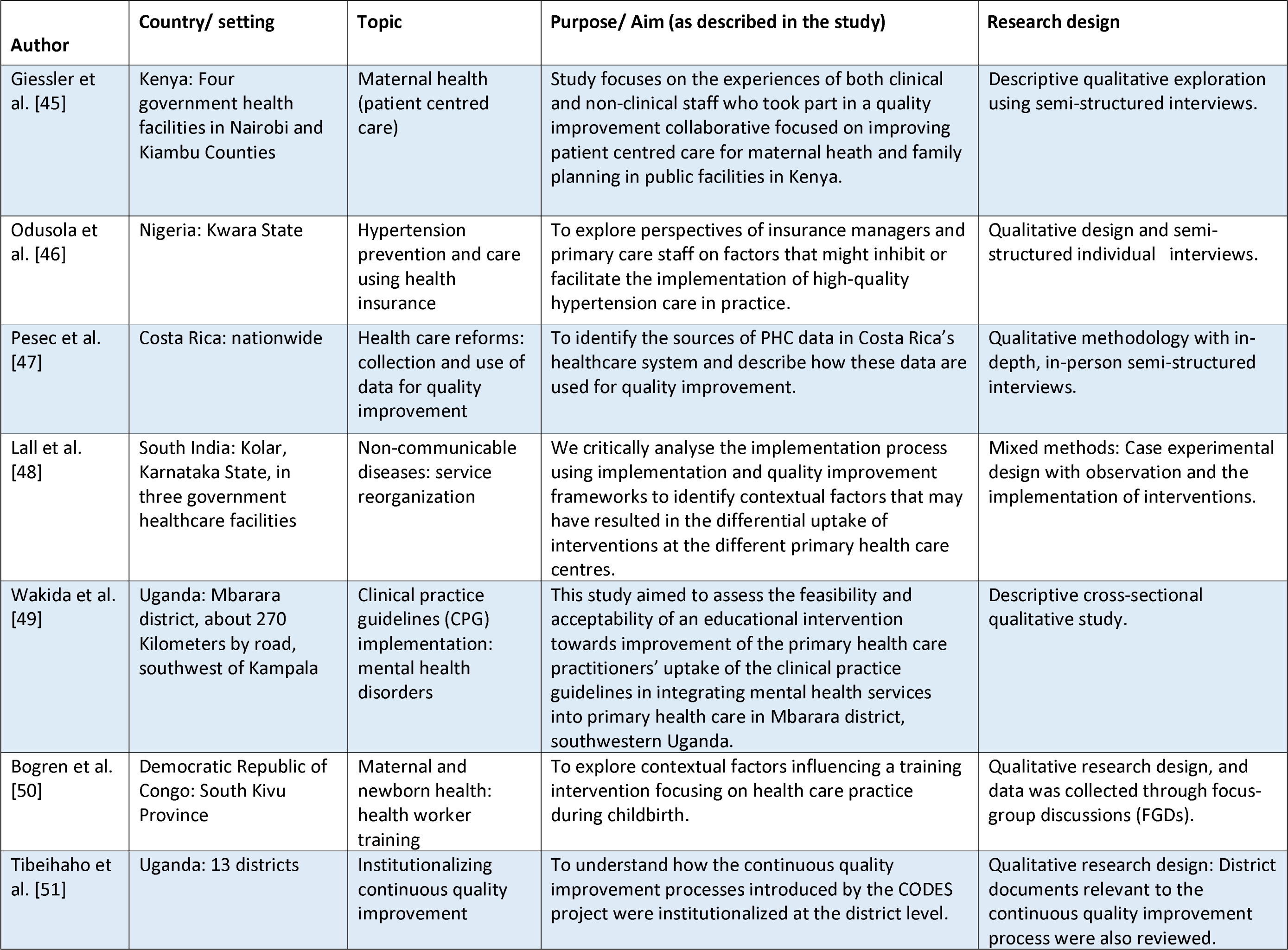
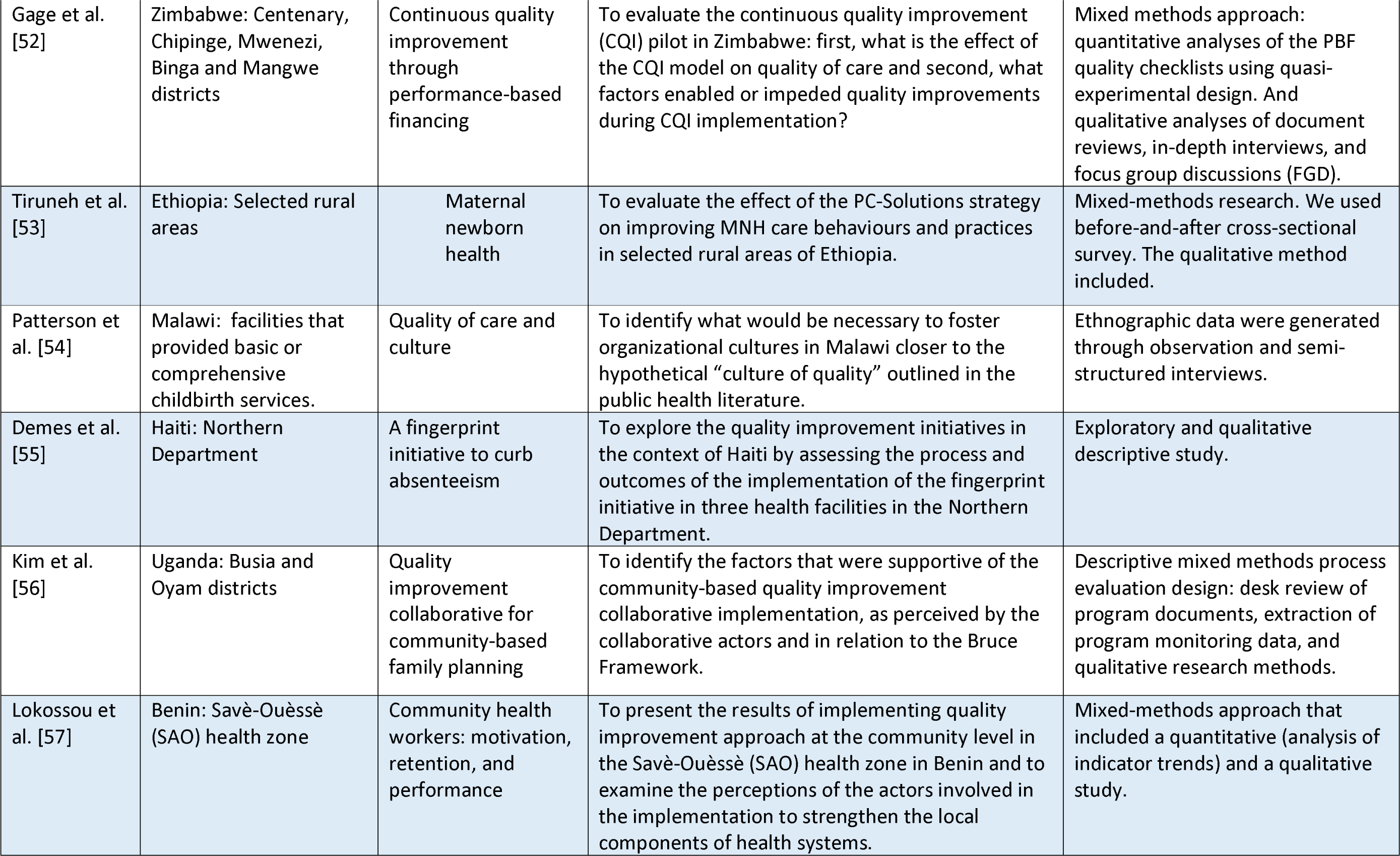
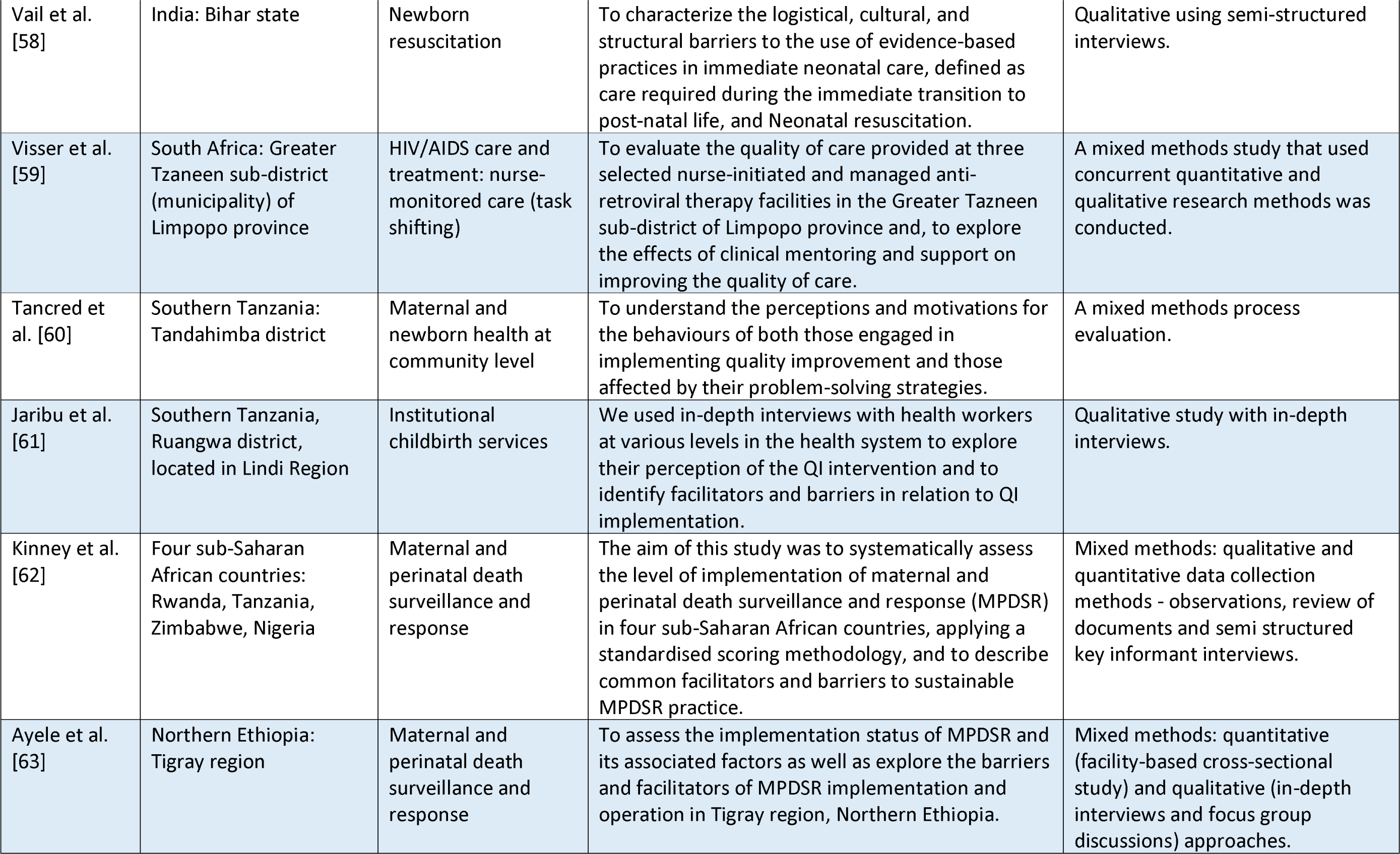
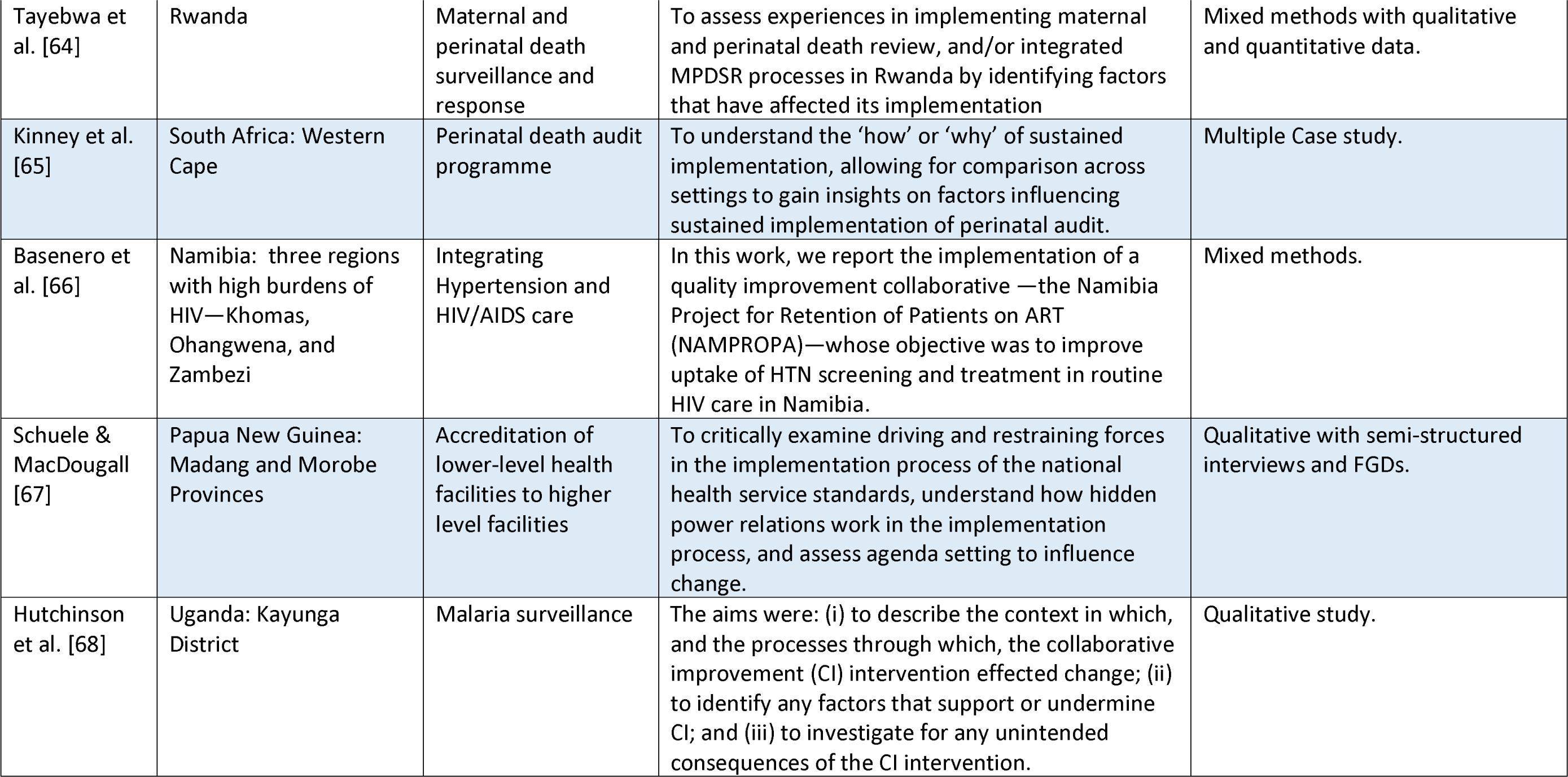
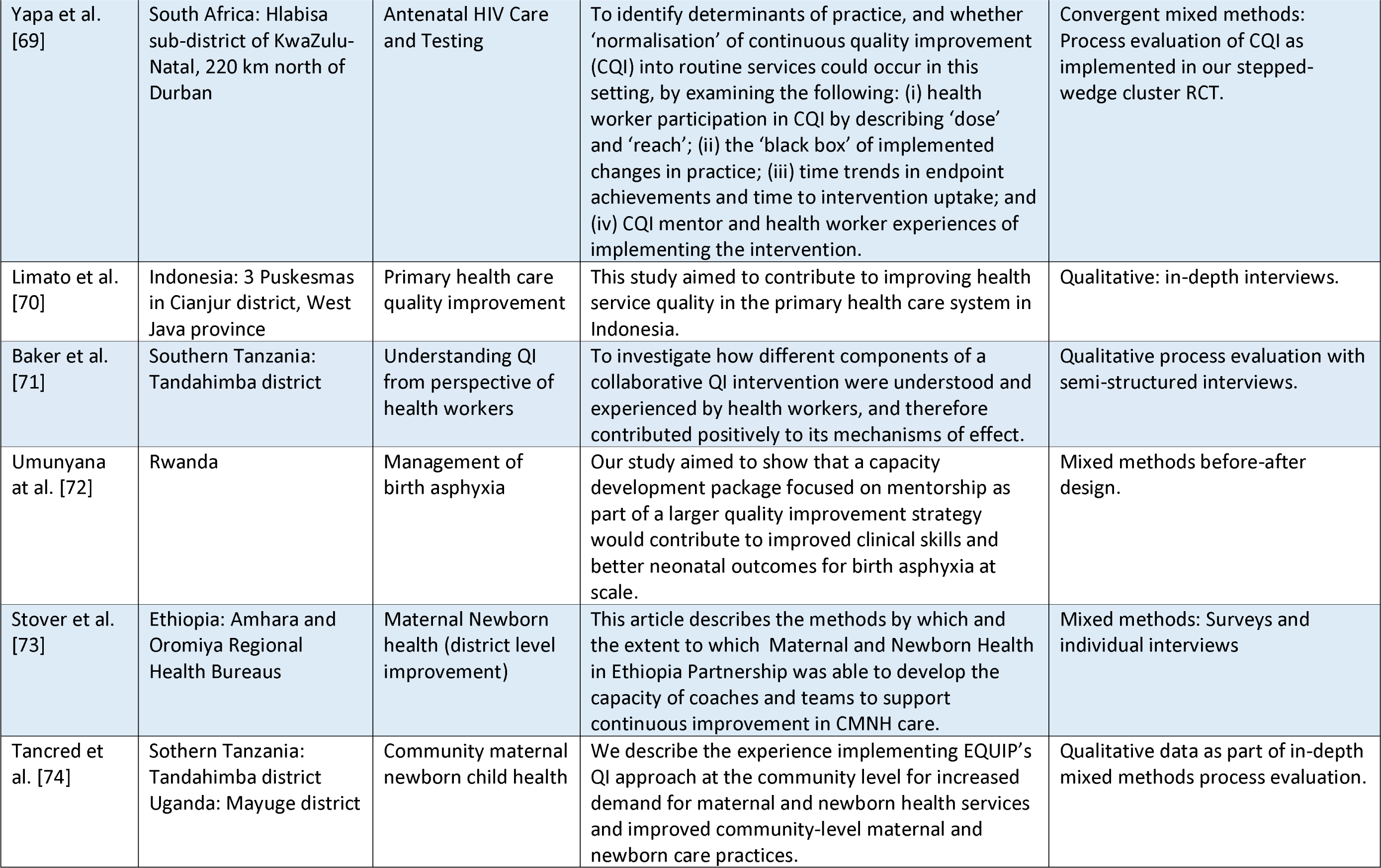
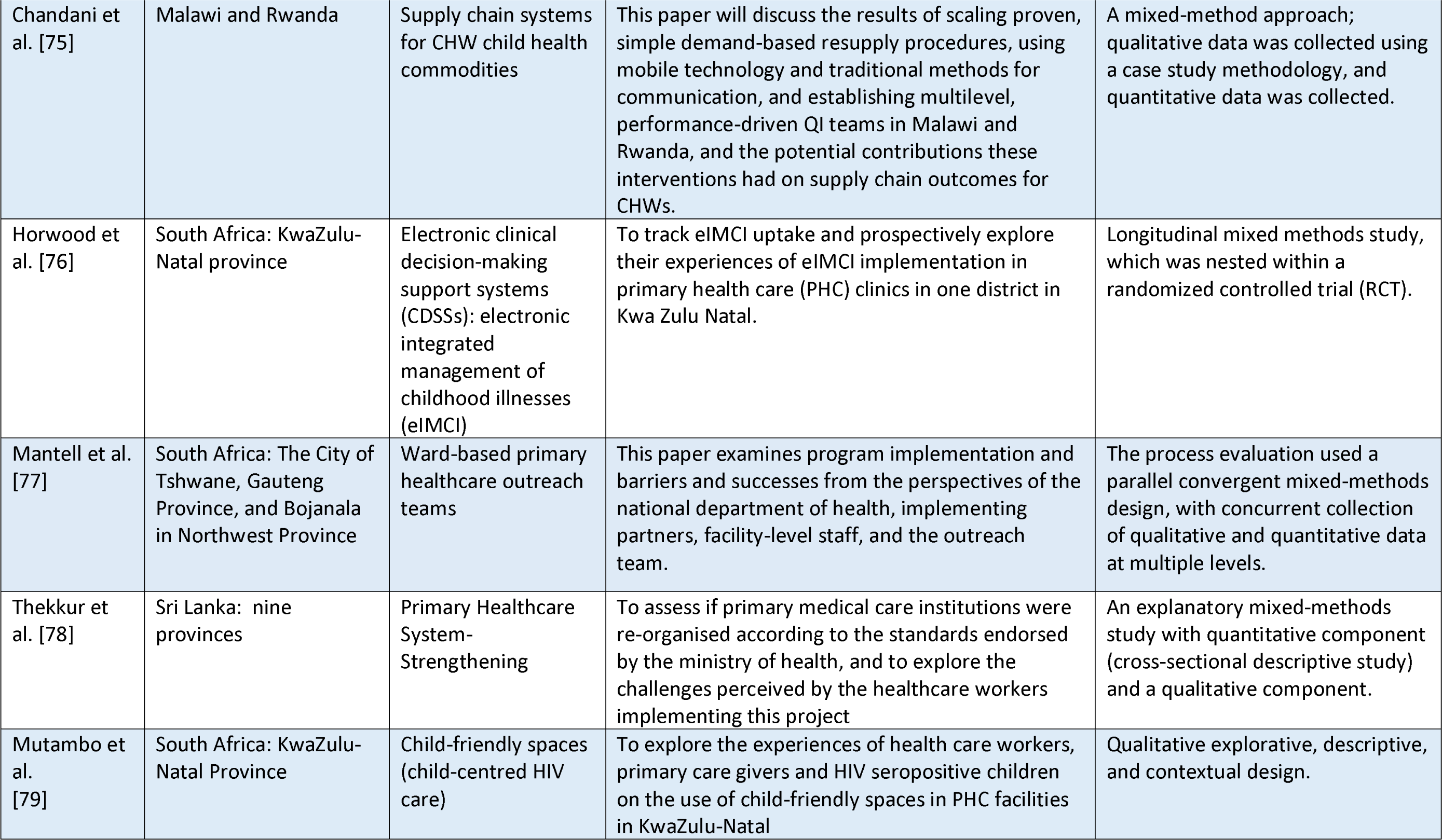
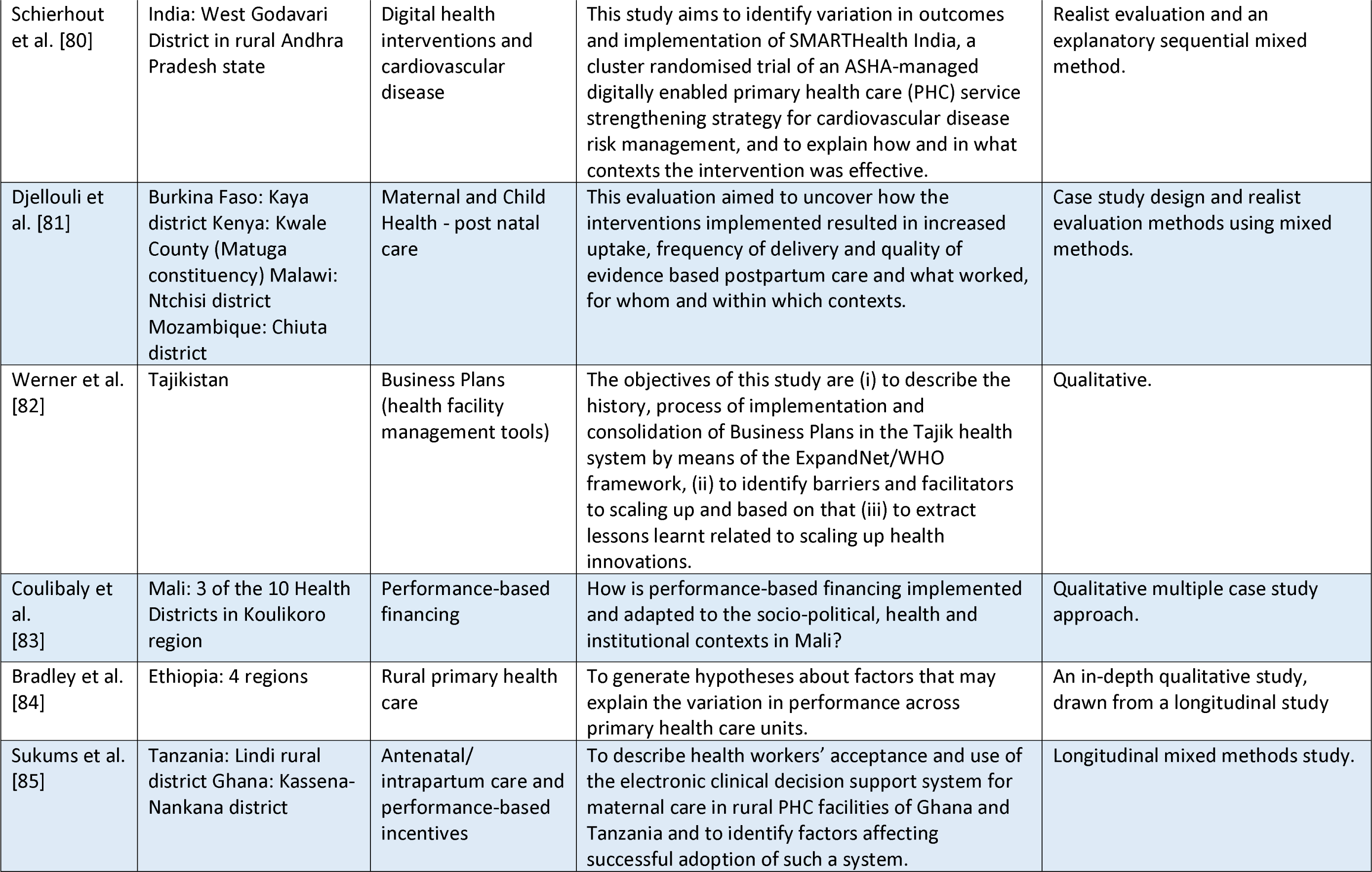
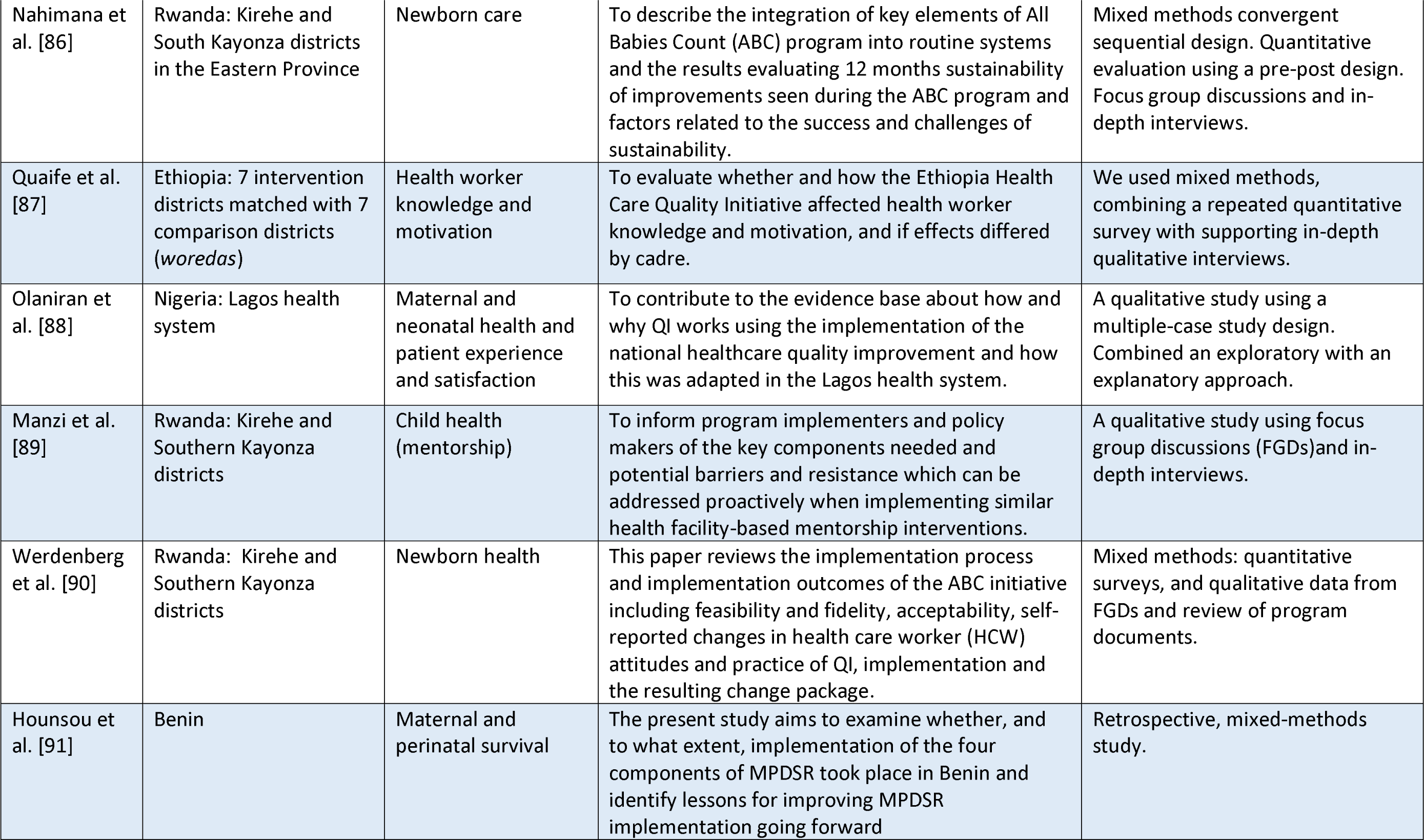
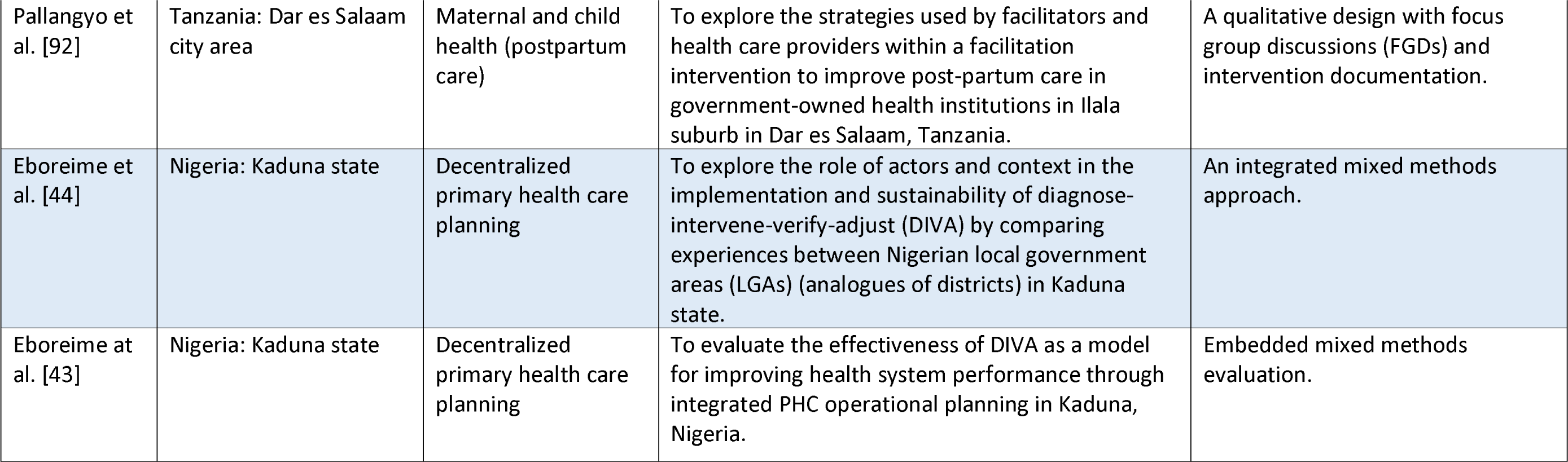
Characteristics of included studies.

### Topics from included research

Researchers overwhelmingly focused on topics related to improvements in maternal and child health (MCH) with twenty-nine studies, including some two conducted in Kenya [45, 81] and some five that focused on maternal and perinatal deaths: in Ethiopia [63]; in Benin [91]; in South Africa [65]; in Rwanda [64] and in Rwanda, Tanzania, Zimbabwe and Nigeria [62]. A summary of studies by topic of focus is contained in supplementary files (S4 Table). As well, five studies documented QI in relation to non-communicable diseases: hypertension service coverage in Nigeria [46]; digital health interventions for cardiovascular health in India [48, 80]; mental health services in Uganda [49]; and integration of hypertension and HIV services in Namibia [66]. Three studies explored QI in HIV/AIDS: nurse-monitored HIV/AIDS care and treatment as part of task-shifting [59] and antenatal HIV care and testing [79] in South Africa, and service expansion through integration in Namibia [66]. One study [68] sought to further the understanding of collaborative QI in malaria surveillance in Uganda. Three studies: in Rwanda and Nigeria [75); in South Africa [76]; and in India [80] investigated the application of digital interventions to improve PHC service delivery.

### Themes

Barriers to and enablers of quality improvement in primary health care at micro, meso- and macro-level were distilled into six themes, guided by the model for understanding success in quality (MUSIQ) and the consolidated framework for implementation research (CFIR), and are described next. Themes are closely related and mutually interacting.

### Theme 1: Microsystem and individual health worker(s) motivation

The willingness and commitment of individual health workers to make improvements, their ability and self-efficacy regarding change efforts, shared values, beliefs, and norms that affect teamwork, interpersonal communication and decision making, and the capacity of health workers and managers to lead QI can constrain or promote QI in PHC settings. Three multi-country studies in Sub-Saharan Africa [62, 81, 85], 19 single country studies in SSA (three apiece in Nigeria, South Africa and Uganda, two each in Benin, Ethiopia and Rwanda, and one study across Benin, Kenya, Mali, Tanzania and Zimbabwe reported various aspects of individual- and microsystem-level barriers to and enablers of QI along with the two studies [47, 55] from Latin America (Costa Rica and Haiti). Studies in Indonesia [70]; in Sri Lanka [78]; in India [48]; in Tajikistan [82]; and in Papua New Guinea [67] also discussed aspects of microsystems and individual health worker motivations for quality improvement. All studies had good quality ratings using the mixed methods appraisal tool.

Health workers and other primary health care stakeholders reported that job satisfaction arising from participating in QI activities was an important source of motivation, encouraging them to increase efforts and stirring up their desire to address the community’s health needs. Added to this, health workers felt extrinsically motivated by financial and non-financial incentives as was the case in Nigeria [46] where Odusola and colleagues found that such inputs bolstered efforts to expand hypertension preventive services and in Haiti where those health workers that perceived an initiative to reduce absenteeism favourably because they thought it promoted openness in the performance-based financing scheme [55]. On the other hand, lack of recognition for putting in effort dimmed motivation levels. Other motivators included a strong desire to help one’s community and appreciation of the justification for a proposed QI project.

Motivation also arose out of observation of positive changes in the PHC setting due to QI and this was underscored by grateful clients or patients. Leadership by PHC facility and district QI mentors who remained committed and were able to showcase the use of context-specific data for QI was also found to enable QI. On the contrary, health workers did not like overlapping QI data streams because this, they perceived, stole time that they would otherwise spend caring for their patients.

Studies also reported the importance of buy-in by health workers and their managers into proposed QI interventions. This was signified by health workers embracing a spirit of personal sacrifice to receive public praise, including by PHC clients from the community. Further, research reports found that participants often embraced QI because they had grown dissatisfied with existing dismal quality of PHC services and felt an urgency to change [67, 76, 81, 83, 89].

Self-efficacy and capability to undertake QI was also highlighted in studies. A high level of technical and managerial proficiency acquired after implementing QI initiatives over time as reported from research in Kenya and Costa Rica [45, 47], promotes effective production, analysis, and use of PHC data for improvement. Moreover, participants in QI felt empowered and competent following training sessions which also served to help develop an understanding of their roles and responsibilities in QI [48] leading to increasing levels of comfort with QI approaches and methods [51]. Health workers reported that they could not spare time to attend QI meetings due to clinical engagements, a possible constraint. Other barriers reported in the literature included the sense of despair with which some easily gave up on QI initiatives when faced with multiple obstacles. An example of this came from a convergent mixed methods process evaluation of continuous quality improvement in South Africa [69] where health workers were discouraged by layers of managerial approval. In such cases across multiple PHC contexts, QI tasks were perceived to be time consuming - reducing health workers’ confidence in the QI initiative - and abandoned [44, 47, 48, 51, 53, 57, 58, 62, 67, 68, 72, 73, 75-77, 83, 85-87, 92].

Health workers developed personal skills through their participation in QI initiatives. Skills such as empathy and enhanced communication with PHC clients reportedly led to deeper connections with fellow health workers but also clients. This was seen to facilitate QI. Still, familiarity with patient-centered approaches to PHC, regular review meetings where gaps and root causes to poor service quality were discussed, and internal supervision where knowledge was shared and additional skills acquired was reported in the literature as important enablers. On the other hand, health workers in PHC who felt inadequately skilled in technical and clinical aspects and in the use of technology reported difficulties engaging effectively in QI [72, 76, 85].

Culture, comprised of shared norms, values, knowledge, artefacts, and practices, was found to play an important role in health workers’ efforts to improve the quality of primary health care (PHC). For example, QI efforts appeared to thrive in PHC settings with strong culture of using data to orchestrate healthcare improvements, where health workers’ attitudes shift to focus more on the needs of patients (e.g., the desire to alleviate pain and reduce suffering), and where HCWs learn better and systematic approaches to solving problems [45, 46, 51, 54]. Additionally, culture of quality manifested in health workers being able to work across disciplinary boundaries, where QI initiatives stir up healthy competition, and where participants reported collective responsibility for cohesion, meritocracy, a strong sense of taking responsibility for failure and success, and high standards in the PHC setting or workplace [54, 63]. Microsystem culture such as working with unsupportive colleagues where workload is not shared and characterized by a rejection of quality checklists [81] was found to be unsupportive of QI. In Indonesia, Limato and others [70] conducted 28 in-depth interviews in West Java Province. This led them to conclude that health workers at government-owned PHC facilities had a general tendency to reject transparency and accountability, which led to the failure of a QI initiative built around performance-based financing. Evidence on workplace culture’s role in boosting or dooming QI interventions also came from other studies in multiple LMIC contexts [47, 53, 58, 59, 62, 64, 65, 68, 76-78, 83].

### Theme 2: Attributes of quality improvement intervention

Component attributes of a quality improvement intervention discussed under this theme include its strength and the quality of evidence underpinning it, how and whether participants perceive it to be beneficial, its cost, potential to be scaled up, and perceived sustainability. Other characteristics of the QI intervention that can enable or constrain its implementation may include its trialability (being trialed in small measures where potential failure is not catastrophic), the ease with which it can be integrated into existing health worker roles and tasks, and whether clients were afforded opportunity to shape its design. Rounding up the key attributes of any QI intervention is the source of the intervention which may dictate its acceptability, its complexity i.e., ease with which implementers understand it, scope, and disruptiveness during roll out; and closely related to this, feasibility (the extent that implementers feel confident that they can adopt the QI intervention) [27, 40].

Sub-Saharan Africa (SSA) studies contributing to this theme included five each from Tanzania and South Africa; four from Ethiopia; and two each from Rwanda and Nigeria. Five African countries (Benin, Kenya, Mali, Namibia and Zimbabwe) each had a single-country study while Tanzania, Rwanda, Kenya, Nigeria Malawi, Burkina Faso, Mozambique, and Ghana were each part of a multi-country study. In Asia, Indonesia, Tajikistan, Sri Lanka and Papua New Guinea each contributed a study with India contributing two. Studies from Haiti and Costa Rica round up the list of those that contribute an understanding of enablers and barriers related to QI intervention attributes in primary health care in LMICs.

Quality improvement implementation is enabled when health workers and managers perceive an intervention to be effective e.g., by observing the desired outcomes for patients and successful acquisition of new skills [43, 62, 67, 71, 73, 76, 77, 83]. A relative advantage accrues when implementers view a new QI initiative as better than current practice and when the intervention is designed to foster collaboration among a diverse team of workers, and even PHC clients. In contrast, QI is constrained when a QI project does not lead to any tangible improvement or is seen to bear negative or unanticipated consequences like creating an administrative burden for already overstretched HCWs that may manifests in multiple reporting channels. Other barriers were reported in the literature: an intervention package that does not envisage nor address other contextual and health systems barriers to successful implementation such as when was QI focused on short term technical fixes but did not address nor consider structural bottlenecks to PHC quality.

Cost, scalability, and sustainability aspects of quality improvement was closely related. As enablers, the design of a QI intervention needs to make provision for long-term work to sustain changes while ensuring that its costs do not overwhelm the PHC system’s capacity [47, 55, 62, 70]. At the same time, QI is scalable when QI interventions are perceived to be easily transferable to a new area of work within a PHC setting, to other health workers, or even to other health facilities by adopting small incremental changes rather than rapid disruptions [43, 51, 56, 73-75]. Additionally, QI interventions are supported by health workers and health facilities when perceived to be sustainable, i.e., when participants feel confident of continued implementation beyond the planned intervention period [44, 47, 49, 55, 65, 69, 73, 77, 82].

The significance of designing QI interventions in a manner that ensures that health workers see alignment between the proposed QI package and their everyday work responsibilities (job expectations in the PHC practice setting) while complementing participants’ and health system’s values was addressed by Ulrike Baker and colleagues [71] in their qualitative process evaluation of quality improvement in Southern Tanzania and Mary Kinney and her counterparts [65] who used multiple case studies to understand sustainability of maternal and perinatal death surveillance (MPDSR) in South Africa. Good examples of facilitating factors regarding trialability pointed to QI interventions that had been adapted and pre-tested to suit local conditions [60, 74]. Barriers that may thwart assimilation included new interventions that are difficult to integrate into routine PHC practice or those that require substantial modifications to service delivery workflows and an array of new skills for practitioners, new initiatives perceived to be inflexible or rigid, in addition to those that do not explicitly build on existing initiatives [43, 44, 80-83, 88, 90, 92].

Paying attention to the preferences of PHC clients when designing QI interventions that affect them was thought to enable QI in addition to health workers’ inputs to intervention design and was outlined by Mutambo and colleagues [79] who explored HCWs’ perspectives during the setting up of child-friendly spaces in PHC clinics in KwaZulu-Natal, South Africa. However, Umunyana and others [72] in Rwanda and Olaniran and colleagues [88] in Nigeria reported that QI interventions that do not allow implementers to make or suggest adaptations might lead to such initiatives being viewed as alien and imposed, potentially leading to their rejection and failure.

Less complex QI interventions focus on a specific problem, are not too general and do not try to address too many things instantly or concurrently. These were some of the enabling factors identified in the literature. Other facilitating factors included having streamlined management structures in their design. Barriers identified by participants in relation to intervention complexity included those that are considered hard to understand, not easily translatable into tangible action plans, and QI interventions perceived as not user-friendly [42, 51, 66-68, 70, 77, 83, 87, 88, 92], and found that QI projects considered feasible, timely and aligned local priorities were widely embraced, contributing to successful implementation.

### Theme 3: Organization and implementing team

Buy-in (ownership), norms and culture, leadership, and decision-making at the organization level complement the tenure, prior experiences, cohesion, and skills of the implementing team to shape QI processes and outcomes. Also, maturity of the organization’s approach to QI, presence of subject matter specialists able and willing to guide health workers at primary health care facilities, and the participation of physicians in QI initiatives received important considerations in research reports included in this review and are described under this theme.

Studies outlined the barriers to and enablers of primary health care quality improvement at the meso level in 15 different countries in Sub-Saharan Africa (SSA) as reported in 36 different articles. The SSA countries include Uganda, Rwanda, Ethiopia, Burkina Faso, Mozambique, and Mali that are low-income settings; lower middle-income countries of Benin, Ghana, Kenya, Malawi, Nigeria, Tanzania and Zimbabwe; and South Africa and Namibia being upper middle-income settings.

Ensuring that leaders, managers, health workers and other stakeholders buy in to QI initiatives in primary health care emerged strongly from the literature. Baker and others [71] found that health care workers (HCWs) were more receptive to continuous quality improvement (CQI) and welcomed on-job-training meant to bolster their skills in Southern Tanzania. This was echoed by Coulibaly and colleagues [83] in Mali where positive reception of a performance-based financing scheme for improving PHC services was noted among the initiative’s strengths. Elsewhere, adequately preparing the team prior to introducing QI, having point persons to champion QI in the health facility and primary health care network, managers and team members who do not mind taking up additional or new responsibilities and an enthusiastic team that readily and publicly commit to PHC quality improvements were also important enablers of QI [45, 46, 49, 51]. In areas where there was little buy in, such as in Papua New Guinea [67] where regional managers exercising their hidden powers opposed QI, and in Indonesia [70] where ‘ego programming’, the tendency by those that perceive themselves to be outside a QI programme to decline participation, QI initiatives faltered. Organizations also rejected QI outright, with some declaring proposed interventions to be unsuitable without due consideration e.g., in Uganda [68] while middle managers in decentralized PHC settings simply went missing and did not cooperate or support frontline HCWs with QI efforts e.g., in Rwanda and Malawi [75].

QI interventions can flourish in organizations and teams with the right norms and where culture is supportive. A new way of solving intractable problems, regular team reviews that are focused on quality of care [51], finding ways to cope positively with scarcity when resources arent adequate and lack of control at lower levels in centralized PHC settings [54] were mentioned. A quality culture with shared values, attitudes, practices at the organization level includes regular data analysis that drives action and improvement cycles, with feedback loops built around effective communication where QI progress is shared with stakeholders who in turn are responsive. Some downsides to quality culture reported in the literature include unchallenged absenteeism by HCWs [55]; decreasing concern for and normalization of common adverse PHC outcomes [58]; adversarial relationships between managers and HCWs; and a perversive lack of accountability where no follow up is done to ascertain achievement of agreed QI work plan targets [62, 78, 81], which constrain the ability of primary health care to meet patient and client needs.

The maturity of an organization in undertaking quality improvement (QI) was reportedly facilitated by accreditation processes which inspire a virtuous cycle of QI. Organizations undergoing accreditation are expected to plan for QI, allocate budgets and subsequently avail resources needed to enhance the quality of PHC services over time [67]. But the presence of concurrent and similar QI programmes in the same organization might introduce fragmentation and bring about confusion regarding organizations’ priorities, a potential barrier [70]. Lack of institutional knowledge, where implementers do not fully understand organizational bureaucracies, can also hamper QI [82] where planned changes are complex and system wide. QI teams with short tenure due to high staff turnover appeared to reduce organizational maturity for QI implementation, e.g., in Benin where QI team members took up new jobs, and lack of community support and irregular monetary incentives affected teams’ longevity [57].

Using pre-post designs with interviews and focus groups, the role of leadership in facilitating QI was reported by Limato and colleagues [70] in Indonesia and Nahimana and colleagues [86] in Rwanda where leaders owned and steered interventions. In contrast, Hounsou and colleagues [91] using mixed methods reported that a lack of interest by managers constrained MPDSR in Benin. Senior leaders, especially, need to actively embrace and publicly show support for QI for it to succeed as health workers do not wish to second guess their bosses’ allegiances [55, 60, 64, 69, 79, 84]. While such champions can drive change within organizations and foster acceptance of QI initiatives, wearing too many hats can contribute to a lack of focus and become a distraction for QI. Weak leadership by governments in LMICs especially means that QI stewardship and monitoring was frequently left to donors and external partners, and this is in part because of lack of clarity in QI leadership arrangements and high turnover of leaders. In a sub-unit in Ethiopia, for example, leadership constantly changed hands [53]. Similarly, Eboreime and colleagues [44] linked weak leadership to organization culture unfovourable for QI, which proved detrimental to efforts to strengthen PHC quality in Kaduna state in Nigeria.

Physician involvement in QI also acted as an enabler and a barrier, depending on the context. Physicians assume leadership and help build other health workers’ skills. However, in Karnataka State in India [48] found QI constrained in situations where the physician over-asserted authority and ignored other team members’ contributions. Findings in Bihar [58], still in India, also highlighted the important gap left when doctors did not take up their roles as QI mentors in the context of management of birth complications for newborns, with fatal consequences.

Positive team experiences from successful legacy QI projects also reportedly produced domino effects e.g., in Tanzania [92] cross-pollination of ideas occurred when successful initiatives were shared across institutions. Incidentally, in both South Africa [65] and Southern India [48] strong social networks among health workers enabled QI whereas less cohesive teams reported worse outcomes. Strong teams also reported better, inclusive decision-making from the start of a QI project and balanced top-down and bottom-up approaches in decision making. Here, diversity was a strength as everyone was involved. A good example came from Uganda [68] where Hutchinson and colleagues used qualitative methods to study collaborative improvement (CI) for malaria surveillance. They report that CI was undertaken by small, committed teams who willingly involved patients and volunteers. Conversely, barriers to QI arise when team leaders do not genuinely involve others like non-technical (auxiliary) staff, who begin to feel sidelined.

Elaborating on the importance of subject matter specialists for advancing QI initiatives, in Uganda [49] participants received excellent support from a mental health specialist who had good knowledge of clinical practice guidelines, joining champions to bolster QI. The development of skills and knowledge also increases when trained team members report back to fellow health care workers, enabling key QI concepts such as Pareto charts, root cause analysis, and PDSA cycles to percolate in the team for a shared understanding [72, 73], with regular on-job training [69]. One-off training that leaves QI team members without adequate knowledge and skills needed to implement QI were characterized as barriers [73].

### Theme 4: Health systems support and capacity

Availability, adequacy, and distribution of resources needed to deliver primary health care (PHC) services to communities were key contextual drivers for QI reported in studies. Studies found weaknesses in primary health care (PHC) systems pillars required for quality enhancements, signifying inadequate capacity for quality improvement. These include gaps in staffing, supplies and commodities, equipment and devices, physical space and infrastructure, data infrastructure and reporting, learning and knowledge systems, management of patient referrals, and leadership and governance. Some enablers of and barriers to QI under this theme e.g., those relating to leadership and management and to staff training and development, inevitably affect and are affected by those discussed in the other themes in this review. Tellingly, no country among the LMICs studied reported adequate or excess levels of resourcing for QI. Consequently, most of this section describes barriers to QI rather than enablers.

Fourty-two studies highlighting various aspects of health systems support and capacity came from 13 different Sub-Saharan Africa countries. There were also five studies conducted in four Asian countries (India, Indonesia, Sri Lanka and Tajikistan) and two studies from Latin America (Haiti and Costa Rica).

The first barrier to QI in LMICs concerns a dearth of health workers which pervades health systems and within these, primary health care delivery structures do not appear exempt. Low numbers, frequent leave of absence, and rapid turnover of staff are each associated with high workload and were reported as important constraints to quality improvement [49, 60, 62-64, 66, 68, 75-78, 81, 85, 86, 90, 94]. Where staff are available, aligning job descriptions and incentives appeared in the literature as a potential enabler of QI.

Adequate, well designed physical space aids intuitive flow of clients, encourages health workers to undertake certain tasks that are important for quality of care such as handwashing or waste segregation, or even providing oversight to acute cases in the newborn unit from the nurses’ station. On the other hand, literature pointed to sub-optimal infrastructure (poorly designed) and or limited physical spaces as barring improvement actions [50, 79, 81, 83, 92]. This manifested as lack of much needed laboratories and pharmacy stores in Sri Lanka (78), for example.

Studies discussed the role of medical equipment and data infrastructure in relation to quality improvement [69, 72, 77-79, 83, 85, 87, 90]. Participatory and data-driven QI activities, revising data and tools to ensure harmonization of reporting systems were found to facilitate QI. Inadequate patient records at the primary health care facility level as well as a lack of equipment, on the other hand, were mentioned as constraining attempts to enhance PHC service delivery and quality. As with equipment and staffing, stockouts of essential supplies and medicines was also reported as barrier to QI in primary health care settings in low- and middle-income countries (LMICs) including but not limited to Sri Lanka [78], India [80], Ethiopia [84], Nigeria and Tanzania [85, 88] and Rwanda [86].

Availability of resources to support quality improvement was the focus of studies in Kenya [45], Uganda (51, 56), Democratic Republic of the Congo [50], Zimbabwe [52], Ethiopia [53, 73, 84], Malawi [54, 75], Haiti (55), India [58], Benin [57, 94], South Africa [59, 65, 69, 77], Tanzania [61, 74, 92], Zimbabwe [62], Rwanda [64], Namibia [66], Indonesia [70], Mozambique and Burkina Faso [81], Tajikistan [82], Mali [83], and Nigeria [43, 44, 88], underscoring its importance to impede QI and shared concerns across many LMIC contexts.

Studies in Rwanda [64, 72], in Namibia [66], in India [80], and (83) in Mali [80] described the need for strong patient referral systems because continuity of care is integral to PHC. Inadequate patient referral systems, they reported, affected QI where the initiative aimed to enhance linkage and networking within a care network. Other enablers uncovered took the form of continuing (medical/health/nursing) education [46] and knowledge exchange platforms [47, 49, 53, 82]. Knowledge exchange platforms, it was reported, could enhance chances of successful quality improvement by breaking down silos and fostering the integration of care packages.

As previously reported under microsystems and QI team and organization support, facilitative and regular follow up and mentorship enabled QI to happen in LMICs. Facilitating aspects such as feedback from the district health management team and mentorship for frontline HCWs supported skills-building and enabled implementers to brainstorm challenges. Unsurprisingly, QI implementing health workers found unpredictable follow up and punitive supervision geared towards fault-finding undesirable for efforts to improve the quality of primary health care.

Quoting program and policy stakeholders in South Africa, Joan Mantell and colleagues [77] cite fragmentation in PHC as a key systems constraint for quality improvement. Also, policies that limit access to PHC budgets as part of larger health systems configuration can also bar QI in LMICs. Conversely, Manisha Yapa and colleagues [69] report that availability of key guidelines and tools, and according to Werner et al. [82], national policies e.g., those that give a high visibility to PHC can indeed foster a supporting environment for PHC-focused quality improvement.

Elsewhere, sub-optimal government policies and guidelines e.g., failure to integrate clinical decision support systems (CDSS) across the entire health system rather than in one or few vertical programmes was a key constraint contributing to non-use by trained health workers [76]. Mutambo and others [79]) also observed that a government policy forbidding the clattering of walls had the unanticipated consequence of limiting the ability of QI implementers to decorate a children’s clinic. The QI team had hoped to encourage play and boost service uptake by making the HIV clinic child friendly. Both studies were conducted in South Africa.

### Theme 5: External environment and structural factors

The external environment forms the larger context in which quality improvement interventions are implemented. It transcends the social, economic, political, legal, and other normative aspects that shape societal and national health systems priorities and may indirectly or directly affect execution of QI projects or initiatives [27, 40]. In the present review, external incentives and societal pressures that drive change, macro-level allocation of resources and other externalities, and community characteristics such as social norms affect QI implementation in varied ways. Such structural factors are not enacted or imposed by social actors intending to shape QI interventions (although they may end up doing just that) but to address other intractable systemic or societal concerns. Thus, it is important for QI implementers, researchers, and policy makers to be aware of these and to make necessary adjustments to their QI programmes, where possible.

Evidence on external environment and structural barriers and enablers that affect QI implementation came from 19 countries reported in 31 studies. Sub-Saharan Africa (SSA) contributed 26 studies from 14 countries while Asia had five countries’ experiences reported in three studies. Six studies were conducted in Rwanda, five in Tanzania, four in South Africa, three in Ethiopia and two each in Malawi and Nigeria. Kenya, Burkina Faso, Mozambique, Namibia, Mali, Benin, Ghana, and Uganda in SSA and Papua New Guinea, Tajikistan, India, Indonesia, and Sri Lanka in Asia each had one research report included in this review. Increased visibility of PHC business plans for donors, high level politicians and citizens in Tajikistan, and its high-level prioritization by the central government, was reportedly an important enabler [82]. On the other hand, studies in Kaduna state in Nigeria [43, 44] reported that the government at state and national level had not prioritized PHC improvements and largely left the implementation of interventions geared towards PHC systems strengthening to donors, placing constraints on the relevant Primary Health Care Development Agency. Interestingly, weak coordination between the central government and semi-autonomous peripheral governments in Tajikistan thwarted the scale up of QI plans due to insufficient intergovernmental engagement [82].

Strong societal norms seep into the health system, through to individual health workers and managers, and shape contexts of health systems where QI is implemented. As an example, Hounsou and colleagues [91] used a retrospective mixed methods approach to explore implementation of the maternal perinatal death surveillance and response (MPDSR) mechanism in Benin and found that a culture of blame had a chilling effect in the reporting and audit of maternal deaths; a similar finding to Ayele et al. [63] in Ethiopia who also used mixed methods with administrative MPDSR data and in-depth interviews to report that health workers feared litigation and blame by relatives of deceased PHC clients. In this context, broad community dissatisfaction with explanations of causes of death and an overly litigatory society. However, in Mali, Coulibaly and colleagues [83] documented positive collaboration among health workers due to strong societal norms that encourage competitiveness, irrespective of place of employment. The inherent competitiveness inspired health workers to put in their best effort in QI implementation.

External pressures and incentives sometimes combined synergistically with socioeconomic policies to enable quality improvement in primary health care. This was the case in Tajikistan where the government introduced, rather serendipitously, a new health financing policy providing for per capita payments for PHC. The policy reduced financial barriers in the provision of PHC services. However, the QI research literature also reported areas where new policies had negative unintended consequences like the introduction of user fees in Rwanda which led to financial difficulties for women seeking ante-natal care, a component of a newly introduced QI package [86]. Expectedly, Wedernberg et al. [90] also reported socio economic challenges for patients that hindered access to PHC services in Rwanda.

Other external issues are more intractable. Impassable or unmotorable roads impede access to PHC clinics for communities and make it difficult for QI supervisors to undertake regular visits. Shaky internet constrains health workers’ from downloading learning materials. And extended power outages make life difficult for both managers and health workers alike. Good telephone connectivity may enable QI by making it easier for mentors to check in with frontline implementers without the necessity of a long, costly road travel. At the same time, good roads make travel within PHC networks easier for both communities and QI teams and supervisors. While responsibility for none of these structural issues lies within the health system, their inadequacies have the effect of introducing bottlenecks in quality improvement efforts, especially in LMICs, where resources are scarce. Added to these, poor weather conditions, unsafe work environments, conflict, and security threats, further complicated matters, and may even see an exodus of skilled health workers besides diverting resources away from life-saving quality PHC. Expanding the list of challenges to QI that was found in the literature is the onset of COVID-19 pandemic which disrupted PHC in Sri Lanka, as was possibly the case globally in early 2020. Multiple research [43, 63, 67, 69, 78, 80, 81, 84, 85] reported these macro level barriers in one form or the other. Nahimana and colleagues [86] add to this long list of protracted constraints detailing how a prolonged drought and famine and the resulting refugee crisis, as happened in in eastern Burundi, rolled back progress in improving PHC in Kirehe district in Rwanda.

### Theme 6: Execution of quality improvement intervention

No quality improvement intervention is going to attain the desired objective unless implemented. Although this theme is being presented last, it is perhaps the most insightful, following this comprehensive synthesis of the evidence on barriers to and enablers of quality improvement in primary health care in low- and middle-income countries. Execution includes elements of dosage and reach, and how the QI intervention is executed (with scope, quality, time, and cost) to achieve the intended results.

The twenty-two studies that underly this theme came from 17 countries. Eighteen of those studies originated from thirteen countries in Sub-Saharan Africa whereas four studies from Asia were derived from four different country contexts. Of the 17 countries in total, six are low-income countries, nine are lower middle-income countries and two are upper middle-income countries. South Africa and Rwanda each had four studies; Ethiopia, Benin, Malawi, and Tanzania each had two studies included and the rest (Mali, Namibia, Papua New Guinea, India, Malawi, Kenya, Mozambique, Burkina Faso, Sri Lanka, Tajikistan, Ghana, and Nigeria) were covered by a single research report.

Dosage (frequency and intensity) and reach (coverage) of QI interventions to a large extent determine whether a QI change package is successful or not. Thus, reaching adequate numbers of implementers with knowledge and skills, whether by offering training sessions repeatedly or targeting and delivering them when most participants are available, were deemed important enablers [69, 70, 71]. Developing results oriented QI work plans and executing these in a participatory manner, ensuring periodic verification of whether a QI intervention is being implemented as planned, using feedback data from PHC facilities, and rolling out a QI package incrementally - where subsequent sessions build on earlier ones in a responsive manner – also facilitated quality improvement [72, 74, 78, 79, 82, 83, 87, 89]. Contrary to these, keeping a limited focus of QI throughout its implementation, not unfurling all planned aspect of an intervention, and late roll of only a few aspects posed major hindrances, signaling a lack of fidelity to the specific QI’s design and intent [44, 81, 94], and its potential failure. This could be attributed to the lack of clear implementation plans, overly ambitious QI work plans, and skewing QI implementation from original plans under pressure from funders, which exacerbate the challenges of QI implementation.

Already described earlier, supervision and mentorship were identified by the health workers among the biggest enablers of QI during the execution stage, according to Umunyana et al. [72]. Baker and colleagues [71] also reported positive impressions of health workers from being visited at their host health facility by mentors and supervisors. However, such visits needed to be reflexive (questioning own stance, habits, values, attitudes) and reflective (learning from everyday experiences) to enable QI. In the case of tech-driven QI such as electronic integrated management of childhood illnesses (eIMCI), promptness with which implementation challenges were addressed also counted as an enabler for improved practice. Non-implementation of support supervision and limited training for implementers was identified as a constraint to QI [76]. When health workers do not practice new skills gained from QI for extended periods, they potentially forget QI techniques, underscoring the importance of ongoing support and mentorship [69, 78, 83]. Being humble and non-judgmental as a mentor-supervisor, Manzi and colleagues [89] reported, was preferred by PHC health workers following interviews and focus group discussion in Rwanda. Such mentors or supervisors assumed a wide range of roles such as facilitators, trainers, coaches, and role models [92] which enabled QI implementation. They could also act as champions, identifying blockers at various levels of the organization early enough and converting them to supporters thereby bolstering QI implementation [63-66, 80, 90].

Engaging communities and targeting multiple stakeholders was further identified in research reports as key enablers, e.g., in Rwanda [64, 86, 90], Tanzania [74], Ethiopia [73, 84], India [80]) and Nigeria [43] besides Burkina Faso, Malawi, Mozambique and Kenya [81], where QI implementers needed to work collaboratively with community resource persons and opinion leaders and make use of local knowledge to tailor their communication. As an enabler, engaging with a diverse array of QI stakeholders during implementation was specifically outlined by Kinney et al. [65] in South Africa, Basenero et al. [66] in Namibia, and Coulibaly et al. [83] in Mali. A boycott of QI by community catchments of primary health care facilities happened in some instances where their local leaders had not been involved in QI implementation, constraining implementation. Also, QI activities geared towards improving access and quality of PHC services were hampered because clients kept off due to previous negative experience when seeking care, and because of limited risk communication by service providers. Nevertheless, reminders in home-based records for patients, where applicable, facilitated good communication between health workers and their clients [62, 63, 65, 66, 80, 83, 90, 92].

Another enabler during QI implementation entailed the redesign of work/patient flows, as described from stakeholders’ experiences in South Africa, Rwanda, Uganda, Ethiopia, and Tanzania. Because sub-optimal physical infrastructure was identified as a key barrier to the provision of quality PHC, QI interventions that sought to re-design the clinic workflow, as needed, in a patient-centered manner, likely made it easier for health workers to adhere to care protocols.

Among others, [63, 88] found that QI implementation is more successful if it includes enhancements in documentation of care processes, and when stocks of key commodities are tracked and reported regularly. Conversely, failure by implementers to keep track of the availability of drugs and other stocks, aside from the actual stockout, constrains implementation. Further, QI roll out should pay due attention to limited staff time and competing tasks as described earlier, which can present significant challenges to participation by HCWs. Failure to consider this may mean that some staff miss numerous QI meetings and training sessions and place avoidable constraints on QI implementation [69].

## Discussion

This review aimed to identify the barriers (constraints) and enablers (facilitators) to quality improvement in primary health care settings of low- and middle-income country contexts. The review supports the notion, overall, that many contextual barriers exist that minimize the effectiveness of quality improvement interventions, initiatives, or projects in these settings. At the same time, the review identified several factors that may promote the implementation of quality improvement interventions in this setting. Barriers and facilitators related to the inherent characteristics of the QI intervention, the immediate (micro) context, the implementing team and host organization at meso level, the larger health systems context, and at macro level, the societal and structural factors. Additional considerations are related to the execution of the QI intervention. These findings are important for those that design, promote, implement, regulate, and sponsor or fund quality improvement. They are also important for users and clients of primary health care services in LMIC countries because they point to how QI interventions can be further enhanced to support the attainment of PHC objectives of equitable, accessible, acceptable, timely, effective, and patient-centered care; and more broadly, health systems and societal development goals.

Reflecting on the review process, one of the challenges faced in selecting studies for inclusion concerned the definition of quality improvement for which there is still no consensus. A second dilemma surfaced around the definition of primary health care – and subsequent isolation of QI interventions in PHC - especially given the interconnectedness of PHC and tertiary (even secondary) care in any given health system. Consequently, decisions had to be made that both optimized sensitivity of the review and minimized selection bias, noting the lack of consensus, especially regarding the definition of quality improvement. The review thus includes studies where actors at the micro, meso and macro levels actively sought to undertake quality improvement for primary health care using diverse approaches. Quality (healthcare) was broadly defined as that which is safe, effective, people-centred, timely, equitable, integrated, and efficient, following the World Health Organization (WHO) guidance [13]. WHO normative guidance plays an outsize role in the formulation of guidelines in LMIC health systems. Accordingly, quality improvement was conceptualized as any deliberate intervention that aimed to enhance any, some or all these aspects of healthcare quality. The definition of primary health care included clinical interventions of curative, rehabilitative and palliative nature, public health interventions meant to improve health at the population level including preventative interventions, and policy level interventions meant to affect health systems domains (financing, human resources, commodities and supplies, infrastructure etc.), if they targeted positive changes in health planning, resourcing, delivery, and outcomes at the district level and below. This inclusive, broad approach makes the review highly relevant to the diverse real-world LMIC contexts in which QI implementation takes place.

The systematic review, in analysing data from included studies, adopted the Model for Understanding Success in Quality, MUSIQ [27] and the Consolidated Framework for Implementation Research, CFIR [40]. This review used concepts and categories from both frameworks to code and later organize the results thematically.

The review found that various barriers and enablers of QI in primary health care in LMIC contexts relate to all the broad categories proposed by MUSIQ and CFIR frameworks, with many being inter-related, reflecting the complexity of health systems in which QI interventions are introduced, implemented, and thereby constrained or enabled. The Miscellaneous category under MUSIQ includes considerations related to the trigger for QI and whether QI tasks are strategic to the organization and were subsumed under the others in the present review.

Accordingly, MUSIQ and CFIR proved useful for organizing the large amount of data derived from 50 diverse studies from equally varied countries and PHC settings. Additions to the CFIR framework [40] further helped with the synthesis and integration.

The results of this review echo those from an earlier umbrella review [26] which included reviews with primary research studies on the effectiveness, performance, and effects of quality management strategies in hospitals. They found 56 reviews focused almost exclusively on South-East Asia, Europe, and North America, with negligible research on the Americas and sub-Saharan Africa (SSA) contexts. Like this present review, Kringos and colleagues found that 35 of the 56 studies frequently reported contextual factors using the MUSIQ framework. The reported barriers and enablers included external environment, organization, QI support and capacity, microsystems, and QI team categories [26].

A more recent realist review [28] explored factors that affect the effectiveness of quality improvement collaboratives (QICs), among the topics covered in the present review. Having synthesized the findings of 32 research abstracts, Zamboni et al [28] reported that factors inherent in external support, QI team, macro or structural aspects of implementation contexts can enable or constrain QICs, resonating with this review.

Like most previously published systematic reviews on quality improvement that have tended to focus only on hospitals, Stokes and colleagues [23] synthesized research on barriers and enablers related to maternity care in LMICs. With a more limited database search covering only MEDLINE and CINAHL, they included nine studies, all of which were based on sub-Saharan Africa. Seven of the studies reviewed by Stokes et al. [23] discussed clinical audits and feedback, like the five in this review that focused on maternal and perinatal death surveillance and response (MPDSR). A key finding of theirs, congruent with this present review, was that intrinsic motivation of health workers was a driver of the implementation of guidelines. However, the present review included community based PHC up to district hospital settings.

This review used an integrative approach [34] with results being synthesized narratively [35]). Studies were found on different topics including malaria surveillance, the application of digital technologies to improve health, expansion of access and quality of HIV/AIDS care, efforts to improve the quality of maternal, newborn and child health services, reduction of childbirth related deaths of women and newborns, and non-communicable diseases. Some studies were cross-cutting and did not look at specific packages of interventions within PHC. These were classified as either continuous quality improvement or quality improvement collaboratives if they explored QI processes using those two approaches, or primary health care systems strengthening, if they were broad enough to include many domains of the health system. These categories helped to compare results and are not necessarily rigid or a priori. Importantly, there are many overlaps among them but this further demonstrates the suitability of the integrative and narrative approaches used for the review, given the review question.

Primary research studies also used many varied approaches to collect and analyse data on constraints and enabler of QI. Key informant interviews, in-depth interviews, semi-structured interviews, document reviews, field notes, participant- and non-participant observations, surveys, focus group and informal discussions, reflexive diaries, and health systems (administrative) performance monitoring were some of the data collection approaches used by QI researchers. Frequently, these were used in combination, with researchers aiming for data saturation. Sample sizes also ranged from a few tens to several hundred for both qualitative and mixed methods design with homogenous and heterogenous groups of quality improvement and primary health care stakeholders. As well, included studies adopted a mix of varied frameworks including MUSIQ, CFIR, Tailored Implementation for Chronic Diseases (TICD), RE-AIM (reach, effectiveness, adoption, implementation, maintenance), COM-B (capability, opportunity, motivation, behaviour), PARIHS (promoting action on research implementation in health services), Breakthrough Model for Improvement, Positive deviance, Data to improvement pathway, and Adaptive management framework. These frameworks, where used, informed QI intervention design, data collection and analysis. Theories were also infused in QI research and included Force Field Analysis derived from Kurt Lewin’s force field theory, Normalization Process Theory (NPT), Barth’s Transactional Model of Culture, Gidden’s Structuration Theory, and Carl May’s Extended NPT. Perhaps given that most QI projects are part of implementation research initiatives, the extended use of frameworks and theories is not surprising. Due to the carefully thought-out theory-driven process evaluations, well-defined samples aiming for data saturation, and method mixing, studies were generally of good quality, having been subjected to critical appraisal, with congruent aims and methods, verifiable findings, and justified conclusions.

### Strength and limitations

The search for literature was comprehensive, covering all major health databases, grey literature repositories, selected websites, and even specialty journals. Moreover, no limiters were applied during search and retrieval. The selection of studies was guided by the review question and definitions adopted a broad and inclusive approach while guarding against scope creep - the tendency for reviews to balloon in size and become unmanageable. Studies were systematically screened and appraised for quality by two reviewers independently. Data extracted from 50 per cent of studies was compared between two independent reviewers for consistency. Together, these measures ensure that the review is relevant, with a low chance of bias, while being applicable across wide LMIC contexts. The review also included studies with a range of methods applicable to process evaluations that elicit contextual barriers to and enablers of quality improvement initiatives in primary health care. This was necessary to answer the review question comprehensively. Of note, the review found relatively recent articles and covered almost all countries in sub-Saharan Africa, seven in Asia and two in Latin America, making it the most comprehensive of its kind so far. Lastly, the use of MUSIQ and CFIR framework that are widely used in reviews and primary research on QI supported rigorous and transparent analysis.

Some limitations exist, nevertheless. Few studies were included from Latin America and Asia, the other continents with many LMIC countries. However, similarities in the contextual barriers to and enablers of QI in PHC irrespective of country context emerged during analysis, and are seemingly shared across LMICs in Africa, Asia, and Latin America. Still, policymakers and practitioners should carefully consider the contexts of included studies before transferring the review’s conclusions to their unique PHC contexts. As there are ongoing debates regarding evolving definitions of QI, some researchers may avoid referring explicitly to QI, and such studies could have been missed. To mitigate this, a broad and inclusive definition that reflects the complex and interconnected nature of social, clinical, and public health interventions in the health system was applied to the review.

## Conclusion

This is the first review of its kind that synthesizes research on quality improvement from low- and middle-income countries with a focus on primary health care. The uncovered themes related to barriers and enablers at the microsystem and individual health worker level, those intrinsic to the QI intervention, others that reside in the organization and team implementing QI, additional ones arising out of the larger health system, external environment including the wider society, and how the QI intervention is executed. The review found many similarities and few contrasts among varied country contexts. Importantly, barriers and enablers are closely related and dynamic, likely affecting and affected by each other. The review found that relatively fewer (22) included studies exploring how the external environment and structural barriers and enablers affect QI implementation. It further found that how QI initiatives are executed had been explored in at least 17 countries out of all the six themes. This signals the opportunity for future research to investigate how wider (macro-level) issues and how the actual implementation process of QI is impeded or promoted to make primary health care better for those that provide, use, fund, regulate or design it in LMIC contexts.

## Supporting information

S1 Checklist. PRISMA checklist

S1 Table. Keywords for database search

S2 Table. Keywords for database search

S3 Fig. MEDLINE EBSCO search strategy

S4 Fig. ProQuest database search strategy

S4 Table. Studies by geographic and income distribution

S5 Table. Themes and sub-themes

S6 Table. QI topic of research

## Supporting information

Fig 1. PRISMA Flow Chart

Supplemental Table 1. Search keywords

Supplemental Figure 3. MEDLINE search

Supplemental Table 4. Studies by geographic and income of country

Supplemental Table 5. Themes and sub-themes

Supplemental Table 6. Topics covered in QI research

Supplemental Figure 4. ProQuest search

## Data Availability

All data produced in the present study i.e., data extracted from primary research reports are available upon reasonable request to the authors.

## Acknowledgements

Professor Mark Limmer provided useful guidance in the development of the research protocol and during the conduct of the systematic review. Dr Tracy Epton of Manchester University also provided useful suggestions regarding the review methodology and approach, which shaped the conduct of this review. John Barbrook provided useful insights into the database search process, from a librarian information scientist perspective, which helped us to fine-tune our literature search strategy. Dr Kevin Oyula undertook quality appraisal and data extraction for one of the included research reports.

