## Supplementary figures and images for "Barriers to and enablers of quality improvement in primary health care in low- and middle-income countries: a systematic review"

### Fig 1. PRISMA Flow Chart

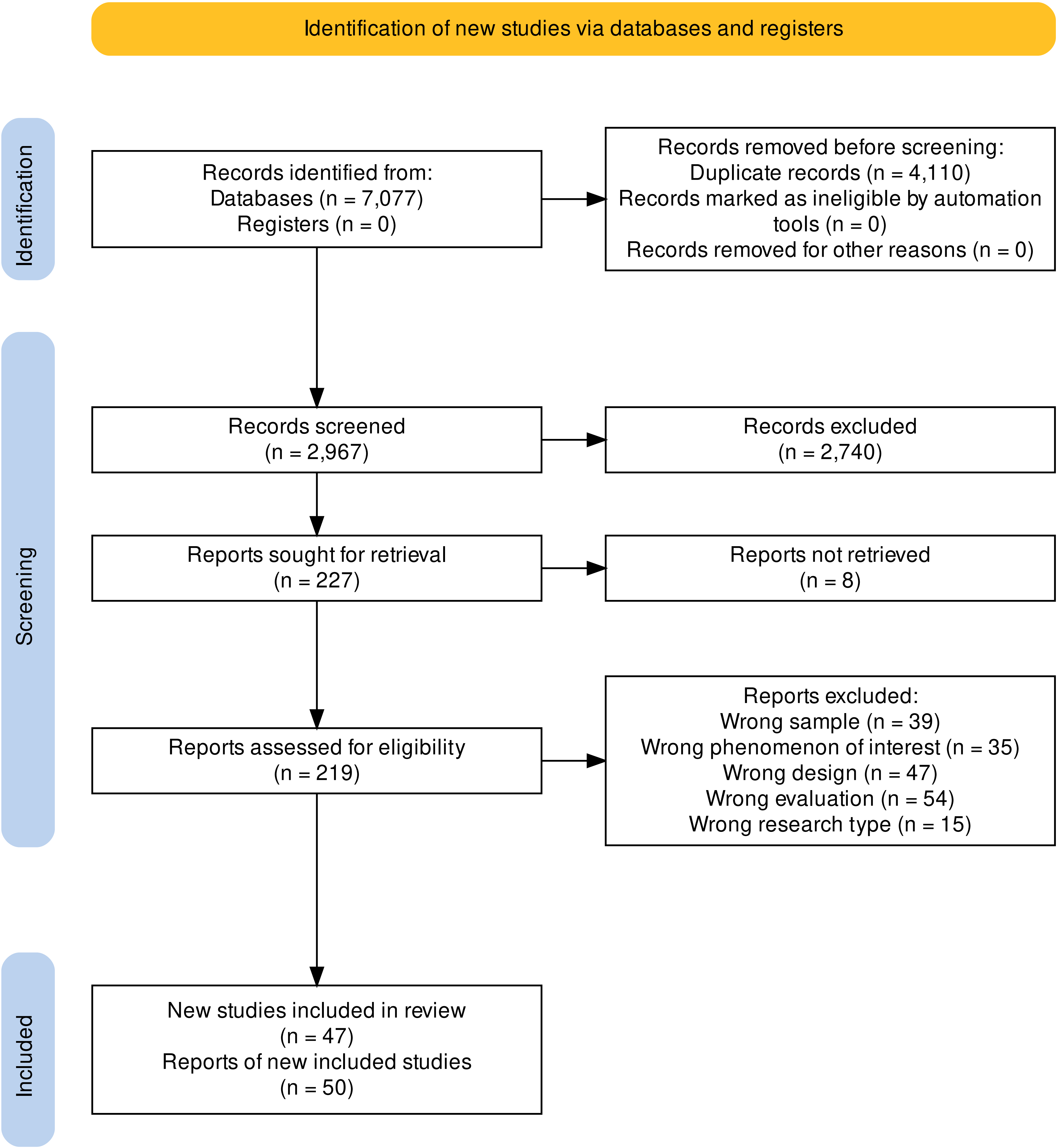
