## Supplemental Table 1. Search keywords for "Barriers to and enablers of quality improvement in primary health care in low- and middle-income countries: a systematic review"

**S1 Table. Key words used when searching databases and websites**

| **Sample size** | **Phenomenon of interest** | **Design of studies** | **Evaluation** | **Research type** |
| --- | --- | --- | --- | --- |
| Health worker (all cadres & levels, stakeholders) | Quality improvement in primary health care | Qualitative OR “Mixed Methods” | Barrier* OR limitation* OR constraint* OR enabler* OR promoter* OR facilitator* OR Attitude* OR belief* OR practice* OR knowledge* OR perception* OR perspective* OR behaviour* OR culture OR motivation OR beliefs OR value* OR factor* | Observation OR Interview OR “Focus Group” OR Survey OR Questionnaire OR “Case Study” OR KII OR IDI OR FGD OR “Participant observation” OR OR “Group Interview” |
| “Health managers” OR  “Quality improvement team” OR  “Quality improvement committee*” OR  “Health service provider” OR  “Primary care team” OR  “Primary care physicians” OR  “Health cent* workers” OR  “Dispensary worker*” OR  “Health post worker*” OR  “Community health worker*” OR “Primary care network” OR Primary Health care network” OR PCN | (“Health care quality improvement” OR “Quality Improvement” OR) AND (Primary Health Care” OR “Essential health care” OR “Basic Health Care” OR QI OR “Quality enhancement” OR (“Curative OR Rehabilitative OR Prevent* OR Promot* AND health) |  |  |  |
