## Supplemental Figure 3. MEDLINE search for "Barriers to and enablers of quality improvement in primary health care in low- and middle-income countries: a systematic review"

MY
EBSCOhost

Wednesday, February 01, 2023 7:50:45 PM

| # | Query | Limiters/Expanders | Last Run Via | Results |
|---|---|---|---|---|
| S12 | AB (reproductive maternal newborn neonatal child adolescent) N5 health AND AB ( afghanistan OR albania OR algeria OR american samoa OR angola OR “antigua and barbuda” OR antigua OR barbuda OR argentina OR armenia OR armenian OR aruba OR azerbaijan OR bahrain OR bangladesh OR barbados OR republic of belarus OR belarus OR byelarus OR belorussia OR byelorussian OR belize OR british honduras OR benin OR dahomey OR bhutan OR bolivia OR “bosnia and herzegovina” OR bosnia OR herzegovina OR botswana OR bechuanaland OR brazil OR brasil OR bulgaria OR burkina faso OR burkina fasso OR upper volta OR burundi OR urundi OR cabo verde OR cape verde OR cambodia OR kampuchea OR khmer republic OR cameroon OR cameron OR cameroun OR central african republic OR ubangi shari OR chad OR chile OR china OR | Limiters - Date of Publication: 20000101-20221231<br>Search modes - SmartText Searching | Interface - EBSCOhost Research Databases<br>Search Screen - Advanced Search<br>Database - MEDLINE Complete | 71 |

colombia OR comoros OR comoro islands OR iles comores OR mayotte OR democratic republic of the congo OR democratic republic congo OR congo OR zaire OR costa rica OR “cote d'ivoire” OR “cote d’ ivoire” OR cote divoire OR cote d ivoire OR ivory coast OR croatia OR cuba OR cyprus OR czech republic OR czechoslovakia OR djibouti OR french somaliland OR dominica OR dominican republic OR ecuador OR egypt OR united arab republic OR el salvador OR equatorial guinea OR spanish guinea OR eritrea OR estonia OR eswatini OR swaziland OR ethiopia OR fiji OR gabon OR gabonese republic OR gambia OR “georgia (republic)” OR georgian OR ghana OR gold coast OR gibraltar OR greece OR grenada OR guam OR guatemala OR guinea OR guinea bissau OR guyana OR british guiana OR haiti OR hispaniola OR honduras OR hungary OR india OR indonesia OR timor OR iran OR iraq OR isle of man OR jamaica OR jordan OR kazakhstan OR kazakh OR kenya OR “democratic people's republic of korea” OR republic of korea OR

north korea OR south
korea OR korea OR
kosovo OR kyrgyzstan
OR kirghizia OR
kirgizstan OR kyrgyz
republic OR kirghiz OR
laos OR lao pdr OR “lao
people's democratic
republic” OR latvia OR
lebanon OR lebanese
republic OR lesotho OR
basutoland OR liberia
OR libya OR libyan arab
jamahiriya OR lithuania
OR macau OR macao
OR republic of north
macedonia OR
macedonia OR
madagascar OR
malagasy republic OR
malawi OR nyasaland
OR malaysia OR malay
federation OR malaya
federation OR maldives
OR indian ocean islands
OR indian ocean OR mali
OR malta OR micronesia
OR federated states of
micronesia OR kiribati
OR marshall islands OR
nauru OR northern
mariana islands OR
palau OR tuvalu OR
mauritania OR mauritius
OR mexico OR moldova
OR moldovian OR
mongolia OR
montenegro OR morocco
OR ifni OR mozambique
OR portuguese east
africa OR myanmar OR
burma OR namibia OR
nepal OR netherlands
antilles OR nicaragua OR
niger OR nigeria OR
oman OR muscat OR
pakistan OR panama OR

papua new guinea OR new guinea OR paraguay OR peru OR philippines OR philipines OR phillipines OR phillippines OR poland OR “polish people's republic” OR portugal OR portuguese republic OR puerto rico OR romania OR russia OR russian federation OR ussr OR soviet union OR union of soviet socialist republics OR rwanda OR ruanda OR samoa OR pacific islands OR polynesia OR samoan islands OR navigator island OR navigator islands OR “sao tome and principe” OR saudi arabia OR senegal OR serbia OR seychelles OR sierra leone OR slovakia OR slovak republic OR slovenia OR melanesia OR solomon island OR solomon islands OR norfolk island OR norfolk islands OR somalia OR south africa OR south sudan OR sri lanka OR ceylon OR “saint kitts and nevis” OR “st. kitts and nevis” OR saint lucia OR “st. lucia” OR “saint vincent and the grenadines” OR saint vincent OR “st. vincent” OR grenadines OR sudan OR suriname OR surinam OR dutch guiana OR netherlands guiana OR syria OR syrian arab republic OR tajikistan OR tadjikistan OR tadzhikistan OR tadzhik OR tanzania OR tanganyika OR thailand OR siam OR timor leste OR east timor OR togo OR togolese republic OR tonga OR “trinidad and tobago” OR trinidad OR tobago OR tunisia OR turkey OR turkmenistan OR turkmen OR uganda OR ukraine OR uruguay OR uzbekistan OR uzbek OR vanuatu OR new hebrides OR venezuela OR vietnam OR viet nam OR middle east OR west bank OR gaza OR palestine OR yemen OR yugoslavia OR zambia OR zimbabwe OR northern rhodesia OR global south OR africa south of the sahara OR sub-saharan africa OR subsaharan africa OR africa, central OR central africa OR africa, northern OR north africa OR northern africa OR magreb OR maghrib OR sahara OR africa, southern OR southern africa OR africa, eastern OR east africa OR eastern africa OR africa, western OR west africa OR western africa OR west indies OR indian ocean islands OR caribbean OR central america OR latin america OR “south and central america” OR south america OR asia, central OR central asia OR asia, northern OR north asia

| | | | | |
|---|---|---|---|---|
| | OR northern asia OR asia, southeastern OR southeastern asia OR south eastern asia OR southeast asia OR south east asia OR asia, western OR western asia OR europe, eastern OR east europe OR eastern europe ) AND ( (Research OR Study) N20 "quality improvement" ) | | | |
| S11 | AB ( "primary health care" OR "primary care" OR "public health care" OR "integrated primary care team" OR "PHC" ) AND AB ( afghanistan OR albania OR algeria OR american samoa OR angola OR “antigua and barbuda” OR antigua OR barbuda OR argentina OR armenia OR armenian OR aruba OR azerbaijan OR bahrain OR bangladesh OR barbados OR republic of belarus OR belarus OR byelarus OR belorussia OR byelorussian OR belize OR british honduras OR benin OR dahomey OR bhutan OR bolivia OR “bosnia and herzegovina” OR bosnia OR herzegovina OR botswana OR bechuanaland OR brazil OR brasil OR bulgaria OR burkina faso OR burkina fasso OR upper volta OR burundi OR urundi OR cabo verde OR cape verde OR | Limiters - Date of Publication: 20000101-20221231<br>Search modes - Boolean/Phrase | Interface - EBSCOhost Research Databases<br>Search Screen - Advanced Search<br>Database - MEDLINE Complete | 63 |

cambodia OR
kampuchea OR khmer
republic OR cameroon
OR cameron OR
cameroun OR central
african republic OR
ubangi shari OR chad
OR chile OR china OR
colombia OR comoros
OR comoro islands OR
iles comores OR mayotte
OR democratic republic
of the congo OR
democratic republic
congo OR congo OR
zaire OR costa rica OR
“cote d'ivoire” OR “cote d’
ivoire” OR cote divoire
OR cote d ivoire OR ivory
coast OR croatia OR
cuba OR cyprus OR
czech republic OR
czechoslovakia OR
djibouti OR french
somaliland OR dominica
OR dominican republic
OR ecuador OR egypt
OR united arab republic
OR el salvador OR
equatorial guinea OR
spanish guinea OR
eritrea OR estonia OR
eswatini OR swaziland
OR ethiopia OR fiji OR
gabon OR gabonese
republic OR gambia OR
“georgia (republic)” OR
georgian OR ghana OR
gold coast OR gibraltar
OR greece OR grenada
OR guam OR guatemala
OR guinea OR guinea
bissau OR guyana OR
british guiana OR haiti
OR hispaniola OR
honduras OR hungary
OR india OR indonesia

OR timor OR iran OR
iraq OR isle of man OR
jamaica OR jordan OR
kazakhstan OR kazakh
OR kenya OR
“democratic people's
republic of korea” OR
republic of korea OR
north korea OR south
korea OR korea OR
kosovo OR kyrgyzstan
OR kirghizia OR
kirgizstan OR kyrgyz
republic OR kirghiz OR
laos OR lao pdr OR “lao
people's democratic
republic” OR latvia OR
lebanon OR lebanese
republic OR lesotho OR
basutoland OR liberia
OR libya OR libyan arab
jamahiriya OR lithuania
OR macau OR macao
OR republic of north
macedonia OR
macedonia OR
madagascar OR
malagasy republic OR
malawi OR nyasaland
OR malaysia OR malay
federation OR malaya
federation OR maldives
OR indian ocean islands
OR indian ocean OR mali
OR malta OR micronesia
OR federated states of
micronesia OR kiribati
OR marshall islands OR
nauru OR northern
mariana islands OR
palau OR tuvalu OR
mauritania OR mauritius
OR mexico OR moldova
OR moldovian OR
mongolia OR
montenegro OR morocco
OR ifni OR mozambique

OR portuguese east africa OR myanmar OR burma OR namibia OR nepal OR netherlands antilles OR nicaragua OR niger OR nigeria OR oman OR muscat OR pakistan OR panama OR papua new guinea OR new guinea OR paraguay OR peru OR philippines OR philipines OR phillipines OR phillippines OR poland OR “polish people's republic” OR portugal OR portuguese republic OR puerto rico OR romania OR russia OR russian federation OR ussr OR soviet union OR union of soviet socialist republics OR rwanda OR ruanda OR samoa OR pacific islands OR polynesia OR samoan islands OR navigator island OR navigator islands OR “sao tome and principe” OR saudi arabia OR senegal OR serbia OR seychelles OR sierra leone OR slovakia OR slovak republic OR slovenia OR melanesia OR solomon island OR solomon islands OR norfolk island OR norfolk islands OR somalia OR south africa OR south sudan OR sri lanka OR ceylon OR “saint kitts and nevis” OR “st. kitts and nevis” OR saint lucia OR “st. lucia” OR “saint vincent and the grenadines” OR saint vincent OR “st. vincent” OR grenadines OR sudan OR suriname OR surinam OR dutch guiana OR netherlands guiana OR syria OR syrian arab republic OR tajikistan OR tadjikistan OR tadzhikistan OR tadzhik OR tanzania OR tanganyika OR thailand OR siam OR timor leste OR east timor OR togo OR togolese republic OR tonga OR “trinidad and tobago” OR trinidad OR tobago OR tunisia OR turkey OR turkmenistan OR turkmen OR uganda OR ukraine OR uruguay OR uzbekistan OR uzbek OR vanuatu OR new hebrides OR venezuela OR vietnam OR viet nam OR middle east OR west bank OR gaza OR palestine OR yemen OR yugoslavia OR zambia OR zimbabwe OR northern rhodesia OR global south OR africa south of the sahara OR sub-saharan africa OR subsaharan africa OR africa, central OR central africa OR africa, northern OR north africa OR northern africa OR magreb OR maghrib OR sahara OR africa, southern OR southern africa OR africa, eastern OR east africa OR eastern africa OR africa, western OR west africa OR western africa OR west indies OR indian

| | | | | |
|---|---|---|---|---|
| | ocean islands OR caribbean OR central america OR latin america OR “south and central america” OR south america OR asia, central OR central asia OR asia, northern OR north asia OR northern asia OR asia, southeastern OR southeastern asia OR south eastern asia OR southeast asia OR south east asia OR asia, western OR western asia OR europe, eastern OR east europe OR eastern europe ) AND ( (Research OR Study) N20 "quality improvement" ) | | | |
| S10 | AB ( "primary health care" OR "primary care" OR "public health care" OR "integrated primary care team" OR "PHC" ) AND AB ( afghanistan OR albania OR algeria OR american samoa OR angola OR “antigua and barbuda” OR antigua OR barbuda OR argentina OR armenia OR armenian OR aruba OR azerbaijan OR bahrain OR bangladesh OR barbados OR republic of belarus OR belarus OR byelarus OR belorussia OR byelorussian OR belize OR british honduras OR benin OR dahomey OR bhutan OR bolivia OR “bosnia and herzegovina” OR bosnia OR herzegovina OR | Limiters - Date of Publication: 20000101-20221231<br>Search modes - Boolean/Phrase | Interface - EBSCOhost Research Databases<br>Search Screen - Advanced Search<br>Database - MEDLINE Complete | 33 |

botswana OR
bechuanaland OR brazil
OR brasil OR bulgaria
OR burkina faso OR
burkina fasso OR upper
volta OR burundi OR
urundi OR cabo verde
OR cape verde OR
cambodia OR
kampuchea OR khmer
republic OR cameroon
OR cameron OR
cameroun OR central
african republic OR
ubangi shari OR chad
OR chile OR china OR
colombia OR comoros
OR comoro islands OR
iles comores OR mayotte
OR democratic republic
of the congo OR
democratic republic
congo OR congo OR
zaire OR costa rica OR
“cote d'ivoire” OR “cote d’
ivoire” OR cote divoire
OR cote d ivoire OR ivory
coast OR croatia OR
cuba OR cyprus OR
czech republic OR
czechoslovakia OR
djibouti OR french
somaliland OR dominica
OR dominican republic
OR ecuador OR egypt
OR united arab republic
OR el salvador OR
equatorial guinea OR
spanish guinea OR
eritrea OR estonia OR
eswatini OR swaziland
OR ethiopia OR fiji OR
gabon OR gabonese
republic OR gambia OR
“georgia (republic)” OR
georgian OR ghana OR
gold coast OR gibraltar

OR greece OR grenada OR guam OR guatemala OR guinea OR guinea bissau OR guyana OR british guiana OR haiti OR hispaniola OR honduras OR hungary OR india OR indonesia OR timor OR iran OR iraq OR isle of man OR jamaica OR jordan OR kazakhstan OR kazakh OR kenya OR “democratic people's republic of korea” OR republic of korea OR north korea OR south korea OR korea OR korea OR kosovo OR kyrgyzstan OR kirghizia OR kirgizstan OR kyrgyz republic OR kirghiz OR laos OR lao pdr OR “lao people's democratic republic” OR latvia OR lebanon OR lebanese republic OR lesotho OR basutoland OR liberia OR libya OR libyan arab jamahiriya OR lithuania OR macau OR macao OR republic of north macedonia OR macedonia OR madagascar OR malagasy republic OR malawi OR nyasaland OR malaysia OR malay federation OR malaya federation OR maldives OR indian ocean islands OR indian ocean OR mali OR malta OR micronesia OR federated states of micronesia OR kiribati OR marshall islands OR nauru OR northern mariana islands OR palau OR tuvalu OR mauritania OR mauritius OR mexico OR moldova OR moldovian OR mongolia OR montenegro OR morocco OR ifni OR mozambique OR portuguese east africa OR myanmar OR burma OR namibia OR nepal OR netherlands antilles OR nicaragua OR niger OR nigeria OR oman OR muscat OR pakistan OR panama OR papua new guinea OR new guinea OR paraguay OR peru OR philippines OR philipines OR phillipines OR phillippines OR poland OR “polish people's republic” OR portugal OR portuguese republic OR puerto rico OR romania OR russia OR russian federation OR ussr OR soviet union OR union of soviet socialist republics OR rwanda OR ruanda OR samoa OR pacific islands OR polynesia OR samoan islands OR navigator island OR navigator islands OR “sao tome and principe” OR saudi arabia OR senegal OR serbia OR seychelles OR sierra leone OR slovakia OR slovak republic OR slovenia OR melanesia OR solomon island OR solomon islands OR norfolk island OR norfolk islands OR somalia OR south africa OR south sudan OR sri lanka OR ceylon OR “saint kitts and nevis” OR “st. kitts and nevis” OR saint lucia OR “st. lucia” OR “saint vincent and the grenadines” OR saint vincent OR “st. vincent” OR grenadines OR sudan OR suriname OR surinam OR dutch guiana OR netherlands guiana OR syria OR syrian arab republic OR tajikistan OR tadjikistan OR tadzhikistan OR tadzhik OR tanzania OR tanganyika OR thailand OR siam OR timor leste OR east timor OR togo OR togolese republic OR tonga OR “trinidad and tobago” OR trinidad OR tobago OR tunisia OR turkey OR turkmenistan OR turkmen OR uganda OR ukraine OR uruguay OR uzbekistan OR uzbek OR vanuatu OR new hebrides OR venezuela OR vietnam OR viet nam OR middle east OR west bank OR gaza OR palestine OR yemen OR yugoslavia OR zambia OR zimbabwe OR northern rhodesia OR global south OR africa south of the sahara OR sub-saharan africa OR subsaharan africa OR africa, central OR central africa OR africa, northern OR north africa OR northern africa OR magreb OR maghrib OR

| | | | | |
|---|---|---|---|---|
| | sahara OR africa, southern OR southern africa OR africa, eastern OR east africa OR eastern africa OR africa, western OR west africa OR western africa OR west indies OR indian ocean islands OR caribbean OR central america OR latin america OR "south and central america" OR south america OR asia, central OR central asia OR asia, northern OR north asia OR northern asia OR asia, southeastern OR southeastern asia OR south eastern asia OR southeast asia OR south east asia OR asia, western OR western asia OR europe, eastern OR east europe OR eastern europe ) AND ( (Research OR Study) N10 "quality improvement" ) | | | |
| S9 | AB ( "primary health care networks* OR MPDSR OR "maternal perinatal death surveillance response" OR "infection prevention control" OR "surgical safety" OR "hand washing" OR "patient safety" ) AND AB ( afghanistan OR albania OR algeria OR american samoa OR angola OR "antigua and barbuda" OR antigua OR barbuda OR argentina OR armenia OR armenian OR aruba OR azerbaijan | Limiters - Date of Publication: 20000101-20221231<br>Search modes - SmartText Searching | Interface - EBSCOhost Research Databases<br>Search Screen - Advanced Search<br>Database - MEDLINE Complete | 15 |

OR bahrain OR
bangladesh OR
barbados OR republic of
belarus OR belarus OR
byelarus OR belorussia
OR byelorussian OR
belize OR british
honduras OR benin OR
dahomey OR bhutan OR
bolivia OR “bosnia and
herzegovina” OR bosnia
OR herzegovina OR
botswana OR
bechuanaland OR brazil
OR brasil OR bulgaria
OR burkina faso OR
burkina fasso OR upper
volta OR burundi OR
urundi OR cabo verde
OR cape verde OR
cambodia OR
kampuchea OR khmer
republic OR cameroon
OR cameron OR
cameroun OR central
african republic OR
ubangi shari OR chad
OR chile OR china OR
colombia OR comoros
OR comoro islands OR
iles comores OR mayotte
OR democratic republic
of the congo OR
democratic republic
congo OR congo OR
zaire OR costa rica OR
“cote d'ivoire” OR “cote d’
ivoire” OR cote divoire
OR cote d ivoire OR ivory
coast OR croatia OR
cuba OR cyprus OR
czech republic OR
czechoslovakia OR
djibouti OR french
somaliland OR dominica
OR dominican republic
OR ecuador OR egypt

OR united arab republic OR el salvador OR equatorial guinea OR spanish guinea OR eritrea OR estonia OR eswatini OR swaziland OR ethiopia OR fiji OR gabon OR gabonese republic OR gambia OR “georgia (republic)” OR georgian OR ghana OR gold coast OR gibraltar OR greece OR grenada OR guam OR guatemala OR guinea OR guinea bissau OR guyana OR british guiana OR haiti OR hispaniola OR honduras OR hungary OR india OR indonesia OR timor OR iran OR iraq OR isle of man OR jamaica OR jordan OR kazakhstan OR kazakh OR kenya OR “democratic people's republic of korea” OR republic of korea OR north korea OR south korea OR korea OR kosovo OR kyrgyzstan OR kirghizia OR kirgizstan OR kyrgyz republic OR kirghiz OR laos OR lao pdr OR “lao people's democratic republic” OR latvia OR lebanon OR lebanese republic OR lesotho OR basutoland OR liberia OR libya OR libyan arab jamahiriya OR lithuania OR macau OR macao OR republic of north macedonia OR macedonia OR madagascar OR

malagasy republic OR
malawi OR nyasaland
OR malaysia OR malay
federation OR malaya
federation OR maldives
OR indian ocean islands
OR indian ocean OR mali
OR malta OR micronesia
OR federated states of
micronesia OR kiribati
OR marshall islands OR
nauru OR northern
mariana islands OR
palau OR tuvalu OR
mauritania OR mauritius
OR mexico OR moldova
OR moldovian OR
mongolia OR
montenegro OR morocco
OR ifni OR mozambique
OR portuguese east
africa OR myanmar OR
burma OR namibia OR
nepal OR netherlands
antilles OR nicaragua OR
niger OR nigeria OR
oman OR muscat OR
pakistan OR panama OR
papua new guinea OR
new guinea OR paraguay
OR peru OR philippines
OR philipines OR
phillipines OR
phillippines OR poland
OR “polish people's
republic” OR portugal OR
portuguese republic OR
puerto rico OR romania
OR russia OR russian
federation OR ussr OR
soviet union OR union of
soviet socialist republics
OR rwanda OR ruanda
OR samoa OR pacific
islands OR polynesia OR
samoan islands OR
navigator island OR

navigator islands OR “sao tome and principe” OR saudi arabia OR senegal OR serbia OR seychelles OR sierra leone OR slovakia OR slovak republic OR slovenia OR melanesia OR solomon island OR solomon islands OR norfolk island OR norfolk islands OR somalia OR south africa OR south sudan OR sri lanka OR ceylon OR “saint kitts and nevis” OR “st. kitts and nevis” OR saint lucia OR “st. lucia” OR “saint vincent and the grenadines” OR saint vincent OR “st. vincent” OR grenadines OR sudan OR suriname OR surinam OR dutch guiana OR netherlands guiana OR syria OR syrian arab republic OR tajikistan OR tadjikistan OR tadzhikistan OR tadzhik OR tanzania OR tanganyika OR thailand OR siam OR timor leste OR east timor OR togo OR togolese republic OR tonga OR “trinidad and tobago” OR trinidad OR tobago OR tunisia OR turkey OR turkmenistan OR turkmen OR uganda OR ukraine OR uruguay OR uzbekistan OR uzbek OR vanuatu OR new hebrides OR venezuela OR vietnam OR viet nam OR middle east OR west bank OR gaza OR palestine OR yemen OR

| | | | | |
|---|---|---|---|---|
| | yugoslavia OR zambia OR zimbabwe OR northern rhodesia OR global south OR africa south of the sahara OR sub-saharan africa OR subsaharan africa OR africa, central OR central africa OR africa, northern OR north africa OR northern africa OR magreb OR maghrib OR sahara OR africa, southern OR southern africa OR africa, eastern OR east africa OR eastern africa OR africa, western OR west africa OR western africa OR west indies OR indian ocean islands OR caribbean OR central america OR latin america OR “south and central america” OR south america OR asia, central OR central asia OR asia, northern OR north asia OR northern asia OR asia, southeastern OR southeastern asia OR south eastern asia OR southeast asia OR south east asia OR asia, western OR western asia OR europe, eastern OR east europe OR eastern europe ) AND ( Research OR Study ) | | | |
| S8 | AB ( "primary health care networks* OR MPDSR OR "maternal perinatal death surveillance response" OR "infection prevention control" OR "surgical safety" OR | Search modes - SmartText Searching | Interface - EBSCOhost Research Databases<br>Search Screen - Advanced Search<br>Database - MEDLINE Complete | 15 |

"hand washing" OR "patient safety" ) AND AB ( afghanistan OR albania OR algeria OR american samoa OR angola OR “antigua and barbuda” OR antigua OR barbuda OR argentina OR armenia OR armenian OR aruba OR azerbaijan OR bahrain OR bangladesh OR barbados OR republic of belarus OR belarus OR byelarus OR belorussia OR byelorussian OR belize OR british honduras OR benin OR dahomey OR bhutan OR bolivia OR “bosnia and herzegovina” OR bosnia OR herzegovina OR botswana OR bechuanaland OR brazil OR brasil OR bulgaria OR burkina faso OR burkina fasso OR upper volta OR burundi OR urundi OR cabo verde OR cape verde OR cambodia OR kampuchea OR khmer republic OR cameroon OR cameron OR cameroun OR central african republic OR ubangi shari OR chad OR chile OR china OR colombia OR comoros OR comoro islands OR iles comores OR mayotte OR democratic republic of the congo OR democratic republic congo OR congo OR zaire OR costa rica OR “cote d'ivoire” OR “cote d’

ivoire” OR cote divoire
OR cote d ivoire OR ivory
coast OR croatia OR
cuba OR cyprus OR
czech republic OR
czechoslovakia OR
djibouti OR french
somaliland OR dominica
OR dominican republic
OR ecuador OR egypt
OR united arab republic
OR el salvador OR
equatorial guinea OR
spanish guinea OR
eritrea OR estonia OR
eswatini OR swaziland
OR ethiopia OR fiji OR
gabon OR gabonese
republic OR gambia OR
“georgia (republic)” OR
georgian OR ghana OR
gold coast OR gibraltar
OR greece OR grenada
OR guam OR guatemala
OR guinea OR guinea
bissau OR guyana OR
british guiana OR haiti
OR hispaniola OR
honduras OR hungary
OR india OR indonesia
OR timor OR iran OR
iraq OR isle of man OR
jamaica OR jordan OR
kazakhstan OR kazakh
OR kenya OR
“democratic people's
republic of korea” OR
republic of korea OR
north korea OR south
korea OR korea OR
kosovo OR kyrgyzstan
OR kirghizia OR
kirgizstan OR kyrgyz
republic OR kirghiz OR
laos OR lao pdr OR “lao
people's democratic
republic” OR latvia OR

lebanon OR lebanese
republic OR lesotho OR
basutoland OR liberia
OR libya OR libyan arab
jamahiriya OR lithuania
OR macau OR macao
OR republic of north
macedonia OR
macedonia OR
madagascar OR
malagasy republic OR
malawi OR nyasaland
OR malaysia OR malay
federation OR malaya
federation OR maldives
OR indian ocean islands
OR indian ocean OR mali
OR malta OR micronesia
OR federated states of
micronesia OR kiribati
OR marshall islands OR
nauru OR northern
mariana islands OR
palau OR tuvalu OR
mauritania OR mauritius
OR mexico OR moldova
OR moldovian OR
mongolia OR
montenegro OR morocco
OR ifni OR mozambique
OR portuguese east
africa OR myanmar OR
burma OR namibia OR
nepal OR netherlands
antilles OR nicaragua OR
niger OR nigeria OR
oman OR muscat OR
pakistan OR panama OR
papua new guinea OR
new guinea OR paraguay
OR peru OR philippines
OR philipines OR
phillipines OR
phillippines OR poland
OR “polish people's
republic” OR portugal OR
portuguese republic OR

puerto rico OR romania OR russia OR russian federation OR ussr OR soviet union OR union of soviet socialist republics OR rwanda OR ruanda OR samoa OR pacific islands OR polynesia OR samoan islands OR navigator island OR navigator islands OR “sao tome and principe” OR saudi arabia OR senegal OR serbia OR seychelles OR sierra leone OR slovakia OR slovak republic OR slovenia OR melanesia OR solomon island OR solomon islands OR norfolk island OR norfolk islands OR somalia OR south africa OR south sudan OR sri lanka OR ceylon OR “saint kitts and nevis” OR “st. kitts and nevis” OR saint lucia OR “st. lucia” OR “saint vincent and the grenadines” OR saint vincent OR “st. vincent” OR grenadines OR sudan OR suriname OR surinam OR dutch guiana OR netherlands guiana OR syria OR syrian arab republic OR tajikistan OR tadjikistan OR tadzhikistan OR tadzhik OR tanzania OR tanganyika OR thailand OR siam OR timor leste OR east timor OR togo OR togolese republic OR tonga OR “trinidad and tobago” OR trinidad OR tobago OR tunisia OR turkey OR turkmenistan OR turkmen OR uganda OR ukraine OR uruguay OR uzbekistan OR uzbek OR vanuatu OR new hebrides OR venezuela OR vietnam OR viet nam OR middle east OR west bank OR gaza OR palestine OR yemen OR yugoslavia OR zambia OR zimbabwe OR northern rhodesia OR global south OR africa south of the sahara OR sub-saharan africa OR subsaharan africa OR africa, central OR central africa OR africa, northern OR north africa OR northern africa OR magreb OR maghrib OR sahara OR africa, southern OR southern africa OR africa, eastern OR east africa OR eastern africa OR africa, western OR west africa OR western africa OR west indies OR indian ocean islands OR caribbean OR central america OR latin america OR “south and central america” OR south america OR asia, central OR central asia OR asia, northern OR north asia OR northern asia OR asia, southeastern OR southeastern asia OR south eastern asia OR southeast asia OR south east asia OR asia, western OR western asia OR europe, eastern OR east europe OR eastern

| | | | | |
|---|---|---|---|---|
| | europe ) AND ( Research OR Study ) | | | |
| S7 | AB ( (“Health care quality improvement” OR “Quality Improvement” OR QI OR “Quality enhancement”) AND (Primary Health Care” OR “Essential health care” OR “Basic Health Care” OR (“Curative OR Rehabilitative OR Prevent* OR Promot* AND health) (Improving Quality OR safety OR effectiveness OR patient-centeredness OR timeliness OR efficiency OR equitable) AND Health* ) AND AB ( afghanistan OR albania OR algeria OR american samoa OR angola OR “antigua and barbuda” OR antigua OR barbuda OR argentina OR armenia OR armenian OR aruba OR azerbaijan OR bahrain OR bangladesh OR barbados OR republic of belarus OR belarus OR byelarus OR belorussia OR byelorussian OR belize OR british honduras OR benin OR dahomey OR bhutan OR bolivia OR “bosnia and herzegovina” OR bosnia OR herzegovina OR botswana OR bechuanaland OR brazil OR brasil OR bulgaria OR burkina faso OR burkina fasso OR upper volta OR burundi OR | Search modes - Find all my search terms | Interface - EBSCOhost Research Databases<br>Search Screen - Advanced Search<br>Database - MEDLINE Complete | 169 |

urundi OR cabo verde
OR cape verde OR
cambodia OR
kampuchea OR khmer
republic OR cameroon
OR cameron OR
cameroun OR central
african republic OR
ubangi shari OR chad
OR chile OR china OR
colombia OR comoros
OR comoro islands OR
iles comores OR mayotte
OR democratic republic
of the congo OR
democratic republic
congo OR congo OR
zaire OR costa rica OR
“cote d'ivoire” OR “cote d’
ivoire” OR cote divoire
OR cote d ivoire OR ivory
coast OR croatia OR
cuba OR cyprus OR
czech republic OR
czechoslovakia OR
djibouti OR french
somaliland OR dominica
OR dominican republic
OR ecuador OR egypt
OR united arab republic
OR el salvador OR
equatorial guinea OR
spanish guinea OR
eritrea OR estonia OR
eswatini OR swaziland
OR ethiopia OR fiji OR
gabon OR gabonese
republic OR gambia OR
“georgia (republic)” OR
georgian OR ghana OR
gold coast OR gibraltar
OR greece OR grenada
OR guam OR guatemala
OR guinea OR guinea
bissau OR guyana OR
british guiana OR haiti
OR hispaniola OR

honduras OR hungary
OR india OR indonesia
OR timor OR iran OR
iraq OR isle of man OR
jamaica OR jordan OR
kazakhstan OR kazakh
OR kenya OR
“democratic people's
republic of korea” OR
republic of korea OR
north korea OR south
korea OR korea OR
kosovo OR kyrgyzstan
OR kirghizia OR
kirgizstan OR kyrgyz
republic OR kirghiz OR
laos OR lao pdr OR “lao
people's democratic
republic” OR latvia OR
lebanon OR lebanese
republic OR lesotho OR
basutoland OR liberia
OR libya OR libyan arab
jamahiriya OR lithuania
OR macau OR macao
OR republic of north
macedonia OR
macedonia OR
madagascar OR
malagasy republic OR
malawi OR nyasaland
OR malaysia OR malay
federation OR malaya
federation OR maldives
OR indian ocean islands
OR indian ocean OR mali
OR malta OR micronesia
OR federated states of
micronesia OR kiribati
OR marshall islands OR
nauru OR northern
mariana islands OR
palau OR tuvalu OR
mauritania OR mauritius
OR mexico OR moldova
OR moldovian OR
mongolia OR

montenegro OR morocco OR ifni OR mozambique OR portuguese east africa OR myanmar OR burma OR namibia OR nepal OR netherlands antilles OR nicaragua OR niger OR nigeria OR oman OR muscat OR pakistan OR panama OR papua new guinea OR new guinea OR paraguay OR peru OR philippines OR philipines OR phillipines OR phillippines OR poland OR “polish people's republic” OR portugal OR portuguese republic OR puerto rico OR romania OR russia OR russian federation OR ussr OR soviet union OR union of soviet socialist republics OR rwanda OR ruanda OR samoa OR pacific islands OR polynesia OR samoan islands OR navigator island OR navigator islands OR “sao tome and principe” OR saudi arabia OR senegal OR serbia OR seychelles OR sierra leone OR slovakia OR slovak republic OR slovenia OR melanesia OR solomon island OR solomon islands OR norfolk island OR norfolk islands OR somalia OR south africa OR south sudan OR sri lanka OR ceylon OR “saint kitts and nevis” OR “st. kitts and nevis” OR saint lucia OR “st. lucia” OR “saint vincent and the grenadines” OR saint vincent OR “st. vincent” OR grenadines OR sudan OR suriname OR surinam OR dutch guiana OR netherlands guiana OR syria OR syrian arab republic OR tajikistan OR tadjikistan OR tadzhikistan OR tadzhik OR tanzania OR tanganyika OR thailand OR siam OR timor leste OR east timor OR togo OR togolese republic OR tonga OR “trinidad and tobago” OR trinidad OR tobago OR tunisia OR turkey OR turkmenistan OR turkmen OR uganda OR ukraine OR uruguay OR uzbekistan OR uzbek OR vanuatu OR new hebrides OR venezuela OR vietnam OR viet nam OR middle east OR west bank OR gaza OR palestine OR yemen OR yugoslavia OR zambia OR zimbabwe OR northern rhodesia OR global south OR africa south of the sahara OR sub-saharan africa OR subsaharan africa OR africa, central OR central africa OR africa, northern OR north africa OR northern africa OR magreb OR maghrib OR sahara OR africa, southern OR southern africa OR africa, eastern OR east africa OR eastern africa OR africa, western OR west africa

| | | | | |
|---|---|---|---|---|
| | OR western africa OR west indies OR indian ocean islands OR caribbean OR central america OR latin america OR "south and central america" OR south america OR asia, central OR central asia OR asia, northern OR north asia OR northern asia OR asia, southeastern OR southeastern asia OR south eastern asia OR southeast asia OR south east asia OR asia, western OR western asia OR europe, eastern OR east europe OR eastern europe ) | | | |
| S6 | AB ( Barrier* OR limitation* OR constraint* OR enabler* OR promoter* OR facilitator* OR Attitude* OR belief* OR practice* OR knowledge* OR perception* OR perspective* OR behaviour* OR culture OR motivation OR beliefs OR value* OR factor* ) AND AB ( afghanistan OR albania OR algeria OR american samoa OR angola OR "antigua and barbuda" OR antigua OR barbuda OR argentina OR armenia OR armenian OR aruba OR azerbaijan OR bahrain OR bangladesh OR barbados OR republic of belarus OR belarus OR byelarus OR belorussia OR byelorussian OR | Search modes - Find all my search terms | Interface - EBSCOhost Research Databases<br>Search Screen - Advanced Search<br>Database - MEDLINE Complete | 405 |

belize OR british
honduras OR benin OR
dahomey OR bhutan OR
bolivia OR “bosnia and
herzegovina” OR bosnia
OR herzegovina OR
botswana OR
bechuanaland OR brazil
OR brasil OR bulgaria
OR burkina faso OR
burkina fasso OR upper
volta OR burundi OR
urundi OR cabo verde
OR cape verde OR
cambodia OR
kampuchea OR khmer
republic OR cameroon
OR cameron OR
cameroun OR central
african republic OR
ubangi shari OR chad
OR chile OR china OR
colombia OR comoros
OR comoro islands OR
iles comores OR mayotte
OR democratic republic
of the congo OR
democratic republic
congo OR congo OR
zaire OR costa rica OR
“cote d'ivoire” OR “cote d’
ivoire” OR cote divoire
OR cote d ivoire OR ivory
coast OR croatia OR
cuba OR cyprus OR
czech republic OR
czechoslovakia OR
djibouti OR french
somaliland OR dominica
OR dominican republic
OR ecuador OR egypt
OR united arab republic
OR el salvador OR
equatorial guinea OR
spanish guinea OR
eritrea OR estonia OR
eswatini OR swaziland

OR ethiopia OR fiji OR gabon OR gabonese republic OR gambia OR “georgia (republic)” OR georgian OR ghana OR gold coast OR gibraltar OR greece OR grenada OR guam OR guatemala OR guinea OR guinea bissau OR guyana OR british guiana OR haiti OR hispaniola OR honduras OR hungary OR india OR indonesia OR timor OR iran OR iraq OR isle of man OR jamaica OR jordan OR kazakhstan OR kazakh OR kenya OR “democratic people's republic of korea” OR republic of korea OR north korea OR south korea OR korea OR kosovo OR kyrgyzstan OR kirghizia OR kirgizstan OR kyrgyz republic OR kirghiz OR laos OR lao pdr OR “lao people's democratic republic” OR latvia OR lebanon OR lebanese republic OR lesotho OR basutoland OR liberia OR libya OR libyan arab jamahiriya OR lithuania OR macau OR macao OR republic of north macedonia OR macedonia OR madagascar OR malagasy republic OR malawi OR nyasaland OR malaysia OR malay federation OR malaya federation OR maldives OR indian ocean islands

OR indian ocean OR mali OR malta OR micronesia OR federated states of micronesia OR kiribati OR marshall islands OR nauru OR northern mariana islands OR palau OR tuvalu OR mauritania OR mauritius OR mexico OR moldova OR moldovian OR mongolia OR montenegro OR morocco OR ifni OR mozambique OR portuguese east africa OR myanmar OR burma OR namibia OR nepal OR netherlands antilles OR nicaragua OR niger OR nigeria OR oman OR muscat OR pakistan OR panama OR papua new guinea OR new guinea OR paraguay OR peru OR philippines OR philipines OR phillipines OR phillippines OR poland OR “polish people's republic” OR portugal OR portuguese republic OR puerto rico OR romania OR russia OR russian federation OR ussr OR soviet union OR union of soviet socialist republics OR rwanda OR ruanda OR samoa OR pacific islands OR polynesia OR samoan islands OR navigator island OR navigator islands OR “sao tome and principe” OR saudi arabia OR senegal OR serbia OR seychelles OR sierra leone OR slovakia OR slovak republic OR slovenia OR melanesia OR solomon island OR solomon islands OR norfolk island OR norfolk islands OR somalia OR south africa OR south sudan OR sri lanka OR ceylon OR “saint kitts and nevis” OR “st. kitts and nevis” OR saint lucia OR “st. lucia” OR “saint vincent and the grenadines” OR saint vincent OR “st. vincent” OR grenadines OR sudan OR suriname OR surinam OR dutch guiana OR netherlands guiana OR syria OR syrian arab republic OR tajikistan OR tadjikistan OR tadzhikistan OR tadzhik OR tanzania OR tanganyika OR thailand OR siam OR timor leste OR east timor OR togo OR togolese republic OR tonga OR “trinidad and tobago” OR trinidad OR tobago OR tunisia OR turkey OR turkmenistan OR turkmen OR uganda OR ukraine OR uruguay OR uzbekistan OR uzbek OR vanuatu OR new hebrides OR venezuela OR vietnam OR viet nam OR middle east OR west bank OR gaza OR palestine OR yemen OR yugoslavia OR zambia OR zimbabwe OR northern rhodesia OR global south OR africa south of the sahara OR sub-saharan africa OR

| | | | | |
|---|---|---|---|---|
| | subsaharan africa OR africa, central OR central africa OR africa, northern OR north africa OR northern africa OR magreb OR maghrib OR sahara OR africa, southern OR southern africa OR africa, eastern OR east africa OR eastern africa OR africa, western OR west africa OR western africa OR west indies OR indian ocean islands OR caribbean OR central america OR latin america OR “south and central america” OR south america OR asia, central OR central asia OR asia, northern OR north asia OR northern asia OR asia, southeastern OR southeastern asia OR south eastern asia OR southeast asia OR south east asia OR asia, western OR western asia OR europe, eastern OR east europe OR eastern europe ) AND AB "quality improvement" N20 health | | | |
| S5 | AB ( Barrier* OR limitation* OR constraint* OR enabler* OR promoter* OR facilitator* OR Attitude* OR belief* OR practice* OR knowledge* OR perception* OR perspective* OR behaviour* OR culture OR motivation OR beliefs OR value* OR factor* ) AND AB ( afghanistan | Search modes - Find all my search terms | Interface - EBSCOhost Research Databases<br>Search Screen - Advanced Search<br>Database - MEDLINE Complete | 285 |

OR albania OR algeria OR american samoa OR angola OR “antigua and barbuda” OR antigua OR barbuda OR argentina OR armenia OR armenian OR aruba OR azerbaijan OR bahrain OR bangladesh OR barbados OR republic of belarus OR belarus OR byelarus OR belorussia OR byelorussian OR belize OR british honduras OR benin OR dahomey OR bhutan OR bolivia OR “bosnia and herzegovina” OR bosnia OR herzegovina OR botswana OR bechuanaland OR brazil OR brasil OR bulgaria OR burkina faso OR burkina fasso OR upper volta OR burundi OR urundi OR cabo verde OR cape verde OR cambodia OR kampuchea OR khmer republic OR cameroon OR cameron OR cameroun OR central african republic OR ubangi shari OR chad OR chile OR china OR colombia OR comoros OR comoro islands OR iles comores OR mayotte OR democratic republic of the congo OR democratic republic congo OR congo OR zaire OR costa rica OR “cote d'ivoire” OR “cote d’ ivoire” OR cote divoire OR cote d ivoire OR ivory coast OR croatia OR

cuba OR cyprus OR
czech republic OR
czechoslovakia OR
djibouti OR french
somaliland OR dominica
OR dominican republic
OR ecuador OR egypt
OR united arab republic
OR el salvador OR
equatorial guinea OR
spanish guinea OR
eritrea OR estonia OR
eswatini OR swaziland
OR ethiopia OR fiji OR
gabon OR gabonese
republic OR gambia OR
“georgia (republic)” OR
georgian OR ghana OR
gold coast OR gibraltar
OR greece OR grenada
OR guam OR guatemala
OR guinea OR guinea
bissau OR guyana OR
british guiana OR haiti
OR hispaniola OR
honduras OR hungary
OR india OR indonesia
OR timor OR iran OR
iraq OR isle of man OR
jamaica OR jordan OR
kazakhstan OR kazakh
OR kenya OR
“democratic people's
republic of korea” OR
republic of korea OR
north korea OR south
korea OR korea OR
kosovo OR kyrgyzstan
OR kirghizia OR
kirgizstan OR kyrgyz
republic OR kirghiz OR
laos OR lao pdr OR “lao
people's democratic
republic” OR latvia OR
lebanon OR lebanese
republic OR lesotho OR
basutoland OR liberia

OR libya OR libyan arab jamahiriya OR lithuania OR macau OR macao OR republic of north macedonia OR macedonia OR madagascar OR malagasy republic OR malawi OR nyasaland OR malaysia OR malay federation OR malaya federation OR maldives OR indian ocean islands OR indian ocean OR mali OR malta OR micronesia OR federated states of micronesia OR kiribati OR marshall islands OR nauru OR northern mariana islands OR palau OR tuvalu OR mauritania OR mauritius OR mexico OR moldova OR moldovian OR mongolia OR montenegro OR morocco OR ifni OR mozambique OR portuguese east africa OR myanmar OR burma OR namibia OR nepal OR netherlands antilles OR nicaragua OR niger OR nigeria OR oman OR muscat OR pakistan OR panama OR papua new guinea OR new guinea OR paraguay OR peru OR philippines OR philipines OR phillipines OR phillippines OR poland OR “polish people's republic” OR portugal OR portuguese republic OR puerto rico OR romania OR russia OR russian federation OR ussr OR soviet union OR union of soviet socialist republics OR rwanda OR ruanda OR samoa OR pacific islands OR polynesia OR samoan islands OR navigator island OR navigator islands OR “sao tome and principe” OR saudi arabia OR senegal OR serbia OR seychelles OR sierra leone OR slovakia OR slovak republic OR slovenia OR melanesia OR solomon island OR solomon islands OR norfolk island OR norfolk islands OR somalia OR south africa OR south sudan OR sri lanka OR ceylon OR “saint kitts and nevis” OR “st. kitts and nevis” OR saint lucia OR “st. lucia” OR “saint vincent and the grenadines” OR saint vincent OR “st. vincent” OR grenadines OR sudan OR suriname OR surinam OR dutch guiana OR netherlands guiana OR syria OR syrian arab republic OR tajikistan OR tadjikistan OR tadzhikistan OR tadzhik OR tanzania OR tanganyika OR thailand OR siam OR timor leste OR east timor OR togo OR togolese republic OR tonga OR “trinidad and tobago” OR trinidad OR tobago OR tunisia OR turkey OR turkmenistan OR turkmen OR uganda OR ukraine OR uruguay

OR uzbekistan OR uzbek OR vanuatu OR new hebrides OR venezuela OR vietnam OR viet nam OR middle east OR west bank OR gaza OR palestine OR yemen OR yugoslavia OR zambia OR zimbabwe OR northern rhodesia OR global south OR africa south of the sahara OR sub-saharan africa OR subsaharan africa OR africa, central OR central africa OR africa, northern OR north africa OR northern africa OR magreb OR maghrib OR sahara OR africa, southern OR southern africa OR africa, eastern OR east africa OR eastern africa OR africa, western OR west africa OR western africa OR west indies OR indian ocean islands OR caribbean OR central america OR latin america OR “south and central america” OR south america OR asia, central OR central asia OR asia, northern OR north asia OR northern asia OR asia, southeastern OR southeastern asia OR south eastern asia OR southeast asia OR south east asia OR asia, western OR western asia OR europe, eastern OR east europe OR eastern europe ) AND AB "quality improvement" N10 health

| | | | | |
|---|---|---|---|---|
| S4 | AB ( Barrier* OR limitation* OR constraint* OR enabler* OR promoter* OR facilitator* OR Attitude* OR belief* OR practice* OR knowledge* OR perception* OR perspective* OR behaviour* OR culture OR motivation OR beliefs OR value* OR factor* ) AND AB ( afghanistan OR albania OR algeria OR american samoa OR angola OR “antigua and barbuda” OR antigua OR barbuda OR argentina OR armenia OR armenian OR aruba OR azerbaijan OR bahrain OR bangladesh OR barbados OR republic of belarus OR belarus OR byelarus OR belorussia OR byelorussian OR belize OR british honduras OR benin OR dahomey OR bhutan OR bolivia OR “bosnia and herzegovina” OR bosnia OR herzegovina OR botswana OR bechuanaland OR brazil OR brasil OR bulgaria OR burkina faso OR burkina fasso OR upper volta OR burundi OR urundi OR cabo verde OR cape verde OR cambodia OR kampuchea OR khmer republic OR cameroon OR cameron OR cameroun OR central african republic OR | Search modes - Find all my search terms | Interface - EBSCOhost Research Databases Search Screen - Advanced Search Database - MEDLINE Complete | 167 |

ubangi shari OR chad
OR chile OR china OR
colombia OR comoros
OR comoro islands OR
iles comores OR mayotte
OR democratic republic
of the congo OR
democratic republic
congo OR congo OR
zaire OR costa rica OR
“cote d'ivoire” OR “cote d’
ivoire” OR cote divoire
OR cote d ivoire OR ivory
coast OR croatia OR
cuba OR cyprus OR
czech republic OR
czechoslovakia OR
djibouti OR french
somaliland OR dominica
OR dominican republic
OR ecuador OR egypt
OR united arab republic
OR el salvador OR
equatorial guinea OR
spanish guinea OR
eritrea OR estonia OR
eswatini OR swaziland
OR ethiopia OR fiji OR
gabon OR gabonese
republic OR gambia OR
“georgia (republic)” OR
georgian OR ghana OR
gold coast OR gibraltar
OR greece OR grenada
OR guam OR guatemala
OR guinea OR guinea
bissau OR guyana OR
british guiana OR haiti
OR hispaniola OR
honduras OR hungary
OR india OR indonesia
OR timor OR iran OR
iraq OR isle of man OR
jamaica OR jordan OR
kazakhstan OR kazakh
OR kenya OR
“democratic people's

republic of korea” OR
republic of korea OR
north korea OR south
korea OR korea OR
kosovo OR kyrgyzstan
OR kirghizia OR
kirgizstan OR kyrgyz
republic OR kirghiz OR
laos OR lao pdr OR “lao
people's democratic
republic” OR latvia OR
lebanon OR lebanese
republic OR lesotho OR
basutoland OR liberia
OR libya OR libyan arab
jamahiriya OR lithuania
OR macau OR macao
OR republic of north
macedonia OR
macedonia OR
madagascar OR
malagasy republic OR
malawi OR nyasaland
OR malaysia OR malay
federation OR malaya
federation OR maldives
OR indian ocean islands
OR indian ocean OR mali
OR malta OR micronesia
OR federated states of
micronesia OR kiribati
OR marshall islands OR
nauru OR northern
mariana islands OR
palau OR tuvalu OR
mauritania OR mauritius
OR mexico OR moldova
OR moldovian OR
mongolia OR
montenegro OR morocco
OR ifni OR mozambique
OR portuguese east
africa OR myanmar OR
burma OR namibia OR
nepal OR netherlands
antilles OR nicaragua OR
niger OR nigeria OR

oman OR muscat OR pakistan OR panama OR papua new guinea OR new guinea OR paraguay OR peru OR philippines OR philipines OR phillipines OR phillippines OR poland OR “polish people's republic” OR portugal OR portuguese republic OR puerto rico OR romania OR russia OR russian federation OR ussr OR soviet union OR union of soviet socialist republics OR rwanda OR ruanda OR samoa OR pacific islands OR polynesia OR samoan islands OR navigator island OR navigator islands OR “sao tome and principe” OR saudi arabia OR senegal OR serbia OR seychelles OR sierra leone OR slovakia OR slovak republic OR slovenia OR melanesia OR solomon island OR solomon islands OR norfolk island OR norfolk islands OR somalia OR south africa OR south sudan OR sri lanka OR ceylon OR “saint kitts and nevis” OR “st. kitts and nevis” OR saint lucia OR “st. lucia” OR “saint vincent and the grenadines” OR saint vincent OR “st. vincent” OR grenadines OR sudan OR suriname OR surinam OR dutch guiana OR netherlands guiana OR syria OR syrian arab republic OR tajikistan OR tadjikistan OR tadzhikistan OR tadzhik OR tanzania OR tanganyika OR thailand OR siam OR timor leste OR east timor OR togo OR togolese republic OR tonga OR “trinidad and tobago” OR trinidad OR tobago OR tunisia OR turkey OR turkmenistan OR turkmen OR uganda OR ukraine OR uruguay OR uzbekistan OR uzbek OR vanuatu OR new hebrides OR venezuela OR vietnam OR viet nam OR middle east OR west bank OR gaza OR palestine OR yemen OR yugoslavia OR zambia OR zimbabwe OR northern rhodesia OR global south OR africa south of the sahara OR sub-saharan africa OR subsaharan africa OR africa, central OR central africa OR africa, northern OR north africa OR northern africa OR magreb OR maghrib OR sahara OR africa, southern OR southern africa OR africa, eastern OR east africa OR eastern africa OR africa, western OR west africa OR western africa OR west indies OR indian ocean islands OR caribbean OR central america OR latin america OR “south and central america” OR south america OR asia, central

| | | | | |
|---|---|---|---|---|
| | OR central asia OR asia, northern OR north asia OR northern asia OR asia, southeastern OR southeastern asia OR south eastern asia OR southeast asia OR south east asia OR asia, western OR western asia OR europe, eastern OR east europe OR eastern europe ) AND AB "quality improvement" N5 health | | | |
| S3 | AB ( survey OR questionnaire OR Observation OR Interview OR “Focus Group” OR Survey OR Questionnaire OR “Case Study” OR KII OR IDI OR FGD OR “Participant observation” OR OR “Group Interview” OR Ethnography OR “Grounded theory” OR phenomenological OR “qualitative content analysis” OR “Realist Evaluation” ) AND AB ( afghanistan OR albania OR algeria OR american samoa OR angola OR “antigua and barbuda” OR antigua OR barbuda OR argentina OR armenia OR armenian OR aruba OR azerbaijan OR bahrain OR bangladesh OR barbados OR republic of belarus OR belarus OR byelarus OR belorussia OR byelorussian OR belize OR british honduras OR benin OR dahomey OR bhutan OR | Search modes - Find all my search terms | Interface - EBSCOhost Research Databases Search Screen - Advanced Search Database - MEDLINE Complete | 107 |

bolivia OR “bosnia and herzegovina” OR bosnia OR herzegovina OR botswana OR bechuanaland OR brazil OR brasil OR bulgaria OR burkina faso OR burkina fasso OR upper volta OR burundi OR urundi OR cabo verde OR cape verde OR cambodia OR kampuchea OR khmer republic OR cameroon OR cameron OR cameroun OR central african republic OR ubangi shari OR chad OR chile OR china OR colombia OR comoros OR comoro islands OR iles comores OR mayotte OR democratic republic of the congo OR democratic republic congo OR congo OR zaire OR costa rica OR “cote d'ivoire” OR “cote d’ ivoire” OR cote divoire OR cote d ivoire OR ivory coast OR croatia OR cuba OR cyprus OR czech republic OR czechoslovakia OR djibouti OR french somaliland OR dominica OR dominican republic OR ecuador OR egypt OR united arab republic OR el salvador OR equatorial guinea OR spanish guinea OR eritrea OR estonia OR eswatini OR swaziland OR ethiopia OR fiji OR gabon OR gabonese republic OR gambia OR

“georgia (republic)” OR
georgian OR ghana OR
gold coast OR gibraltar
OR greece OR grenada
OR guam OR guatemala
OR guinea OR guinea
bissau OR guyana OR
british guiana OR haiti
OR hispaniola OR
honduras OR hungary
OR india OR indonesia
OR timor OR iran OR
iraq OR isle of man OR
jamaica OR jordan OR
kazakhstan OR kazakh
OR kenya OR
“democratic people's
republic of korea” OR
republic of korea OR
north korea OR south
korea OR korea OR
kosovo OR kyrgyzstan
OR kirghizia OR
kirgizstan OR kyrgyz
republic OR kirghiz OR
laos OR lao pdr OR “lao
people's democratic
republic” OR latvia OR
lebanon OR lebanese
republic OR lesotho OR
basutoland OR liberia
OR libya OR libyan arab
jamahiriya OR lithuania
OR macau OR macao
OR republic of north
macedonia OR
macedonia OR
madagascar OR
malagasy republic OR
malawi OR nyasaland
OR malaysia OR malay
federation OR malaya
federation OR maldives
OR indian ocean islands
OR indian ocean OR mali
OR malta OR micronesia
OR federated states of

micronesia OR kiribati
OR marshall islands OR
nauru OR northern
mariana islands OR
palau OR tuvalu OR
mauritania OR mauritius
OR mexico OR moldova
OR moldovian OR
mongolia OR
montenegro OR morocco
OR ifni OR mozambique
OR portuguese east
africa OR myanmar OR
burma OR namibia OR
nepal OR netherlands
antilles OR nicaragua OR
niger OR nigeria OR
oman OR muscat OR
pakistan OR panama OR
papua new guinea OR
new guinea OR paraguay
OR peru OR philippines
OR philipines OR
phillipines OR
phillippines OR poland
OR “polish people's
republic” OR portugal OR
portuguese republic OR
puerto rico OR romania
OR russia OR russian
federation OR ussr OR
soviet union OR union of
soviet socialist republics
OR rwanda OR ruanda
OR samoa OR pacific
islands OR polynesia OR
samoan islands OR
navigator island OR
navigator islands OR
“sao tome and principe”
OR saudi arabia OR
senegal OR serbia OR
seychelles OR sierra
leone OR slovakia OR
slovak republic OR
slovenia OR melanesia
OR solomon island OR

solomon islands OR norfolk island OR norfolk islands OR somalia OR south africa OR south sudan OR sri lanka OR ceylon OR “saint kitts and nevis” OR “st. kitts and nevis” OR saint lucia OR “st. lucia” OR “saint vincent and the grenadines” OR saint vincent OR “st. vincent” OR grenadines OR sudan OR suriname OR surinam OR dutch guiana OR netherlands guiana OR syria OR syrian arab republic OR tajikistan OR tadjikistan OR tadzhikistan OR tadzhik OR tanzania OR tanganyika OR thailand OR siam OR timor leste OR east timor OR togo OR togolese republic OR tonga OR “trinidad and tobago” OR trinidad OR tobago OR tunisia OR turkey OR turkmenistan OR turkmen OR uganda OR ukraine OR uruguay OR uzbekistan OR uzbek OR vanuatu OR new hebrides OR venezuela OR vietnam OR viet nam OR middle east OR west bank OR gaza OR palestine OR yemen OR yugoslavia OR zambia OR zimbabwe OR northern rhodesia OR global south OR africa south of the sahara OR sub-saharan africa OR subsaharan africa OR africa, central OR central africa OR africa, northern

| | | | | |
|---|---|---|---|---|
| | OR north africa OR northern africa OR magreb OR maghrib OR sahara OR africa, southern OR southern africa OR africa, eastern OR east africa OR eastern africa OR africa, western OR west africa OR western africa OR west indies OR indian ocean islands OR caribbean OR central america OR latin america OR “south and central america” OR south america OR asia, central OR central asia OR asia, northern OR north asia OR northern asia OR asia, southeastern OR southeastern asia OR south eastern asia OR southeast asia OR south east asia OR asia, western OR western asia OR europe, eastern OR east europe OR eastern europe ) AND AB "quality improvement" N5 health | | | |
| S2 | AB ( "qual* research" OR "mixed methods" ) AND AB ( afghanistan OR albania OR algeria OR american samoa OR angola OR “antigua and barbuda” OR antigua OR barbuda OR argentina OR armenia OR armenian OR aruba OR azerbaijan OR bahrain OR bangladesh OR barbados OR republic of belarus OR belarus OR byelarus OR belorussia OR byelorussian OR | Search modes - Find all my search terms | Interface - EBSCOhost Research Databases<br>Search Screen - Advanced Search<br>Database - MEDLINE Complete | 323 |

belize OR british honduras OR benin OR dahomey OR bhutan OR bolivia OR “bosnia and herzegovina” OR bosnia OR herzegovina OR botswana OR bechuanaland OR brazil OR brasil OR bulgaria OR burkina faso OR burkina fasso OR upper volta OR burundi OR urundi OR cabo verde OR cape verde OR cambodia OR kampuchea OR khmer republic OR cameroon OR cameron OR cameroun OR central african republic OR ubangi shari OR chad OR chile OR china OR colombia OR comoros OR comoro islands OR iles comores OR mayotte OR democratic republic of the congo OR democratic republic congo OR congo OR zaire OR costa rica OR “cote d'ivoire” OR “cote d’ ivoire” OR cote divoire OR cote d ivoire OR ivory coast OR croatia OR cuba OR cyprus OR czech republic OR czechoslovakia OR djibouti OR french somaliland OR dominica OR dominican republic OR ecuador OR egypt OR united arab republic OR el salvador OR equatorial guinea OR spanish guinea OR eritrea OR estonia OR eswatini OR swaziland

OR ethiopia OR fiji OR gabon OR gabonese republic OR gambia OR “georgia (republic)” OR georgian OR ghana OR gold coast OR gibraltar OR greece OR grenada OR guam OR guatemala OR guinea OR guinea bissau OR guyana OR british guiana OR haiti OR hispaniola OR honduras OR hungary OR india OR indonesia OR timor OR iran OR iraq OR isle of man OR jamaica OR jordan OR kazakhstan OR kazakh OR kenya OR “democratic people's republic of korea” OR republic of korea OR north korea OR south korea OR korea OR kosovo OR kyrgyzstan OR kirghizia OR kirgizstan OR kyrgyz republic OR kirghiz OR laos OR lao pdr OR “lao people's democratic republic” OR latvia OR lebanon OR lebanese republic OR lesotho OR basutoland OR liberia OR libya OR libyan arab jamahiriya OR lithuania OR macau OR macao OR republic of north macedonia OR macedonia OR madagascar OR malagasy republic OR malawi OR nyasaland OR malaysia OR malay federation OR malaya federation OR maldives OR indian ocean islands

OR indian ocean OR mali OR malta OR micronesia OR federated states of micronesia OR kiribati OR marshall islands OR nauru OR northern mariana islands OR palau OR tuvalu OR mauritania OR mauritius OR mexico OR moldova OR moldovian OR mongolia OR montenegro OR morocco OR ifni OR mozambique OR portuguese east africa OR myanmar OR burma OR namibia OR nepal OR netherlands antilles OR nicaragua OR niger OR nigeria OR oman OR muscat OR pakistan OR panama OR papua new guinea OR new guinea OR paraguay OR peru OR philippines OR philipines OR phillipines OR phillippines OR poland OR “polish people's republic” OR portugal OR portuguese republic OR puerto rico OR romania OR russia OR russian federation OR ussr OR soviet union OR union of soviet socialist republics OR rwanda OR ruanda OR samoa OR pacific islands OR polynesia OR samoan islands OR navigator island OR navigator islands OR “sao tome and principe” OR saudi arabia OR senegal OR serbia OR seychelles OR sierra leone OR slovakia OR slovak republic OR slovenia OR melanesia OR solomon island OR solomon islands OR norfolk island OR norfolk islands OR somalia OR south africa OR south sudan OR sri lanka OR ceylon OR “saint kitts and nevis” OR “st. kitts and nevis” OR saint lucia OR “st. lucia” OR “saint vincent and the grenadines” OR saint vincent OR “st. vincent” OR grenadines OR sudan OR suriname OR surinam OR dutch guiana OR netherlands guiana OR syria OR syrian arab republic OR tajikistan OR tadjikistan OR tadzhikistan OR tadzhik OR tanzania OR tanganyika OR thailand OR siam OR timor leste OR east timor OR togo OR togolese republic OR tonga OR “trinidad and tobago” OR trinidad OR tobago OR tunisia OR turkey OR turkmenistan OR turkmen OR uganda OR ukraine OR uruguay OR uzbekistan OR uzbek OR vanuatu OR new hebrides OR venezuela OR vietnam OR viet nam OR middle east OR west bank OR gaza OR palestine OR yemen OR yugoslavia OR zambia OR zimbabwe OR northern rhodesia OR global south OR africa south of the sahara OR sub-saharan africa OR

| | | | | |
|---|---|---|---|---|
| | subsaharan africa OR africa, central OR central africa OR africa, northern OR north africa OR northern africa OR magreb OR maghrib OR sahara OR africa, southern OR southern africa OR africa, eastern OR east africa OR eastern africa OR africa, western OR west africa OR western africa OR west indies OR indian ocean islands OR caribbean OR central america OR latin america OR “south and central america” OR south america OR asia, central OR central asia OR asia, northern OR north asia OR northern asia OR asia, southeastern OR southeastern asia OR south eastern asia OR southeast asia OR south east asia OR asia, western OR western asia OR europe, eastern OR east europe OR eastern europe ) AND AB "primary health*" | | | |
| S1 | AB ( "Quality Improvement+" OR "Quality of Health Care+" OR "Total Quality Management" OR "Quality Assurance, Health Care" OR "Quality Indicators, Health Care+" ) AND AB ( afghanistan OR albania OR algeria OR american samoa OR angola OR “antigua and barbuda” OR antigua OR | Search modes - Find all my search terms | Interface - EBSCOhost Research Databases Search Screen - Advanced Search Database - MEDLINE Complete | 43 |

barbuda OR argentina OR armenia OR armenian OR aruba OR azerbaijan OR bahrain OR bangladesh OR barbados OR republic of belarus OR belarus OR byelarus OR belorussia OR byelorussian OR belize OR british honduras OR benin OR dahomey OR bhutan OR bolivia OR “bosnia and herzegovina” OR bosnia OR herzegovina OR botswana OR bechuanaland OR brazil OR brasil OR bulgaria OR burkina faso OR burkina fasso OR upper volta OR burundi OR urundi OR cabo verde OR cape verde OR cambodia OR kampuchea OR khmer republic OR cameroon OR cameron OR cameroun OR central african republic OR ubangi shari OR chad OR chile OR china OR colombia OR comoros OR comoro islands OR iles comores OR mayotte OR democratic republic of the congo OR democratic republic congo OR congo OR zaire OR costa rica OR “cote d'ivoire” OR “cote d’ ivoire” OR cote divoire OR cote d ivoire OR ivory coast OR croatia OR cuba OR cyprus OR czech republic OR czechoslovakia OR djibouti OR french somaliland OR dominica OR dominican republic OR ecuador OR egypt OR united arab republic OR el salvador OR equatorial guinea OR spanish guinea OR eritrea OR estonia OR eswatini OR swaziland OR ethiopia OR fiji OR gabon OR gabonese republic OR gambia OR “georgia (republic)” OR georgian OR ghana OR gold coast OR gibraltar OR greece OR grenada OR guam OR guatemala OR guinea OR guinea bissau OR guyana OR british guiana OR haiti OR hispaniola OR honduras OR hungary OR india OR indonesia OR timor OR iran OR iraq OR isle of man OR jamaica OR jordan OR kazakhstan OR kazakh OR kenya OR “democratic people's republic of korea” OR republic of korea OR north korea OR south korea OR korea OR kosovo OR kyrgyzstan OR kirghizia OR kirgizstan OR kyrgyz republic OR kirghiz OR laos OR lao pdr OR “lao people's democratic republic” OR latvia OR lebanon OR lebanese republic OR lesotho OR basutoland OR liberia OR libya OR libyan arab jamahiriya OR lithuania OR macau OR macao OR republic of north

macedonia OR
macedonia OR
madagascar OR
malagasy republic OR
malawi OR nyasaland
OR malaysia OR malay
federation OR malaya
federation OR maldives
OR indian ocean islands
OR indian ocean OR mali
OR malta OR micronesia
OR federated states of
micronesia OR kiribati
OR marshall islands OR
nauru OR northern
mariana islands OR
palau OR tuvalu OR
mauritania OR mauritius
OR mexico OR moldova
OR moldovian OR
mongolia OR
montenegro OR morocco
OR ifni OR mozambique
OR portuguese east
africa OR myanmar OR
burma OR namibia OR
nepal OR netherlands
antilles OR nicaragua OR
niger OR nigeria OR
oman OR muscat OR
pakistan OR panama OR
papua new guinea OR
new guinea OR paraguay
OR peru OR philippines
OR philipines OR
phillipines OR
phillippines OR poland
OR “polish people's
republic” OR portugal OR
portuguese republic OR
puerto rico OR romania
OR russia OR russian
federation OR ussr OR
soviet union OR union of
soviet socialist republics
OR rwanda OR ruanda
OR samoa OR pacific

islands OR polynesia OR samoan islands OR navigator island OR navigator islands OR “sao tome and principe” OR saudi arabia OR senegal OR serbia OR seychelles OR sierra leone OR slovakia OR slovak republic OR slovenia OR melanesia OR solomon island OR solomon islands OR norfolk island OR norfolk islands OR somalia OR south africa OR south sudan OR sri lanka OR ceylon OR “saint kitts and nevis” OR “st. kitts and nevis” OR saint lucia OR “st. lucia” OR “saint vincent and the grenadines” OR saint vincent OR “st. vincent” OR grenadines OR sudan OR suriname OR surinam OR dutch guiana OR netherlands guiana OR syria OR syrian arab republic OR tajikistan OR tadjikistan OR tadzhikistan OR tadzhik OR tanzania OR tanganyika OR thailand OR siam OR timor leste OR east timor OR togo OR togolese republic OR tonga OR “trinidad and tobago” OR trinidad OR tobago OR tunisia OR turkey OR turkmenistan OR turkmen OR uganda OR ukraine OR uruguay OR uzbekistan OR uzbek OR vanuatu OR new hebrides OR venezuela OR vietnam OR viet nam

OR middle east OR west bank OR gaza OR palestine OR yemen OR yugoslavia OR zambia OR zimbabwe OR northern rhodesia OR global south OR africa south of the sahara OR sub-saharan africa OR subsaharan africa OR africa, central OR central africa OR africa, northern OR north africa OR northern africa OR magreb OR maghrib OR sahara OR africa, southern OR southern africa OR africa, eastern OR east africa OR eastern africa OR africa, western OR west africa OR western africa OR west indies OR indian ocean islands OR caribbean OR central america OR latin america OR “south and central america” OR south america OR asia, central OR central asia OR asia, northern OR north asia OR northern asia OR asia, southeastern OR southeastern asia OR south eastern asia OR southeast asia OR south east asia OR asia, western OR western asia OR europe, eastern OR east europe OR eastern europe )
