## Supplemental Table 4. Studies by geographic and income of country for "Barriers to and enablers of quality improvement in primary health care in low- and middle-income countries: a systematic review"

**S4 Table. Geographic focus of included studies by country income status**

| **Country Income Classification** | **Geographic region** | | |
| --- | --- | --- | --- |
|  | **Sub-Saharan Africa** | **Asia** | **Latin America** |
| Low-income | Wakida et al. (2019)- Uganda; Bogren et al. (2021)- DRC; Tibeihaho et al (2021)- Uganda; Tiruneh et al. (2020) - Ethiopia; Kim et al. (2019)- Uganda; Ayele et al. (2019)- Ethiopia; Tayebwa et al. (2020)- Rwanda; Hutchinson et al. (2021)- Uganda;; Umunyana et al. (2020)- Rwanda; Stover et al. (2014)- Ethiopia; Djellouli et al. (2016)- Kenya, Malawi, Burkina Faso and Mozambique; Coulibaly et al. (2020)- Mali; Bradley et al. (2012)- Ethiopia; Nahimana et al. (2021)- Rwanda; Quaife et al. (2021)- Ethiopia; Manzi et al. (2014)- Rwanda; Werdenberg et al. (2018)- Rwanda; Kinney et al (2020)- Tanzania, Nigeria, Rwanda and Zimbabwe | none | Demes et al. (2021) |
| Lower-middle income | Gage et al. (2021)- Zimbabwe; Giessler et al. (2020)- Kenya; Odusola et al. (2016)- Nigeria; Sukums et al. (2015)- Tanzania and Ghana; Olaniran et al. (2022)- Nigeria; Eboreime et al. (2018)- Nigeria; Kinney et al (2020) – Tanzania, Nigeria, Rwanda and Zimbabwe; Djellouli et al. (2016)- Kenya, Malawi, Burkina Faso and Mozambique; Patterson et al. (2021)- Malawi; Lokossou et al. (2019)- Benin; Tancred et al (2017)- Tanzania; Jaribu et al (2016)- Tanzania; Baker et al. (2018)- Tanzania; Tancred et al. (2018)- Tanzania; Chandani et al. (2017)- Malawi and Nigeria; Hounsou et al. (2022)- Benin; Pallangyo et al. (2018)- Tanzania; Sukums et al. (2015)- Tanzania and Ghana | Lall et al. (2020)- India; Vail et al. (2018)- India; Schuele & MacDougall (2022)- Papua New Guinea; Limato et al. (2019)- Indonesia; Schierhout et al. (2021)- India; Werner et al. (2021)- Tajikistan; Thekkur et al (2022)- Sri Lanka | none |
| Upper-middle income | Visser et al. (2018)- South Africa; Kinney et al. (2022)- South Africa; Basenero et al. (2022)- Namibia; Yapa et al. (2022)- South Africa; Horwood et al. (2023)- South Africa; Mantell et al. (2022)- South Africa; Mutambo, Shumba and Hlongwana (2020)- South Africa |  | Pesec et al. (2021) |
