## Supplemental Table 5. Themes and sub-themes for "Barriers to and enablers of quality improvement in primary health care in low- and middle-income countries: a systematic review"

**S5 Table. Analytical framework with themes and sub-themes**

| **Theme** | **Sub-themes** | **Study - country** |
| --- | --- | --- |
| Microsystem: individual health worker motivation for quality improvement | *Enablers:*  -developing empathy and better communication with clients  -Intrinsic motivation i.e., job satisfaction from participation in QI activities motivates health workers to put in more effort and strong desire to help one’s own community  -increased familiarity with patient-centered care approaches, deeper connections between health worker and clients  -extrinsic motivation drawn from financial incentives and understanding rationale for QI  -strong culture of valuing data as a tool to drive improvements  -high level of technical and managerial proficiency promotes effective data collection, analysis, and use gained over time  -feeling empowered and competent after participating in training  -better understanding of roles and responsibilities in QI by health workers and increasing levels of comfort with QI tools  -personal motivation after observing changes due to QI and being thanked by clients/ patients  -regular review meeting to identify gaps and root causes, action planning to address gaps  -health workers inspired by committed health facility/district leaders and QI mentors  -health workers shift attitude to focus more on patient needs with desire to alleviate pain and suffering and reduce deaths  - health workers learn and embrace better ways of solving problems and become more systematic, working across disciplinary boundaries  -district managers’ ability to use contextualized data for QI  -health workers like internal supervision for knowledge sharing and skills development  -QI intervention promotes transparency and stirs up healthy competition  -NGO-owned health facility worker’s norms embrace accountability (performance-driven)  -embrace of personal sacrifice and effort to earn public praise for health workers  -growing dissatisfaction with poor state of service quality  -shared values such as cohesion, merit, individual responsibility, maintaining high standards of work  *Barriers:*  -no spare time for health worker to attend QI meetings due to clinical duties  -financial disincentives lead to frustration and waning interest in QI  -overlapping data systems increase distract from provision of care to patients  -public (government-owned) health facilities reject QI focused on greater transparency and accountability due ingrained  -sensing despair and easily giving up on QI initiatives  -self-efficacy is limited when more manager approvals are needed to carry out work tasks than are necessary and staff feel unskilled (technical/clinical areas and ICT)  -tasks perceived to be time-consuming lower health worker confidence  -unsupportive colleagues at the workplace  -lack of recognition of presumed hard work  -negative culture that rejects use of care delivery checklists and declines referrals even when indicated | **Africa (Low-income):** Tibeihaho et al (2021) – Uganda; Kim et al (2019) - Uganda; Hutchinson et al (2021) - Uganda; Gage et al (2022) - Zimbabwe; Baker et al (2018) - Tanzania; Coulibaly et al (2020) - Mali; Lokossou et al (2019) - Benin; Stover et al (2014) - Ethiopia; Quaife et al (2021) - Ethiopia; Manzi et al (2014) - Rwanda; Werdenberg et al (2018) - Rwanda; Hounsou et al (2022), Benin  **Africa (Lower middle-income):** Giessler et al (2020) - Kenya; Eboreime et al (2018) - Nigeria; Eboreime et al (2019) - Nigeria; Olaniran et al (2022) Nigeria; Odusola et al (2016) - Nigeria  **Africa (Upper middle-income)**: Yapa et al (2022) - South Africa; Horwood et al (2023) - South Africa; Kinney et al (2022) – South Africa  **Asia (UMIC):** Limato et al. (2019) - Indonesia; Thekkur et al (2022) - Sri Lanka; Lall et al (2020) - India; Werner et al (2021) - Tajikistan; Schuele and MacDougall (2022) - Papua New Guinea  **Americas (LIC):** Demes et al (2021) - Haiti  **Americas (UMIC):** Pesec et al (2021) - Costa Rica  **Multi-country:** Djellouli et al (2016) - Malawi, Kenya, Burkina Faso and Mozambique; Kinney et al (2020) -Tanzania, Nigeria, Rwanda, Zimbabwe; Sukums et al (2015) - Tanzania and Ghana |
| QI Intervention Attributes | *Enablers:*  -QI project implementation perceived to be effective i.e., positive outcomes for patients and health workers (implementers) also acquire new skills and knowledge  -QI project is considered feasible, timely and well aligned local priorities  -health workers see a high degree of fit between QI package, their job responsibilities and practice expectations  -health workers see a relative advantage of QI package versus current practice  -QI intervention adapted and pre-tested to suit local implementation conditions  -Intervention is focused on a specific problem, is not too general and does not try to address too many things at once  -participants feel confident continuing with QI even post-intervention period  -QI intervention can be scaled up to other areas, health facilities, or health workers in need  -QI project details clear management structures and does not ignore or assume this  -project design fosters collaboration among diverse workers and even clients  -Intervention design incorporates and complements participants/health system’s values  -QI intervention design makes provision for long-term work to sustain changes and its costs do not overwhelm the systems’ resource capacity  -Intervention adopts small incremental changes informed by feedback mechanisms rather than big rapid leaps  -intervention design incorporates client preferences, not only health workers’ ideas  *Barriers:*  -QI project does not lead to any observable improvements  -QI implementation plans do not attain targeted levels of penetration (low does/reach)  -QI intervention package is hard to understand, not easy to translate into tangible action points, and perceived as not user-friendly  -lack of clear implementation plan for QI intervention  -QI intervention is difficult to integrate in routine practice and or requires substantial modifications to workflows and additional new skills  -in technology-driven QI, perception that the new approach is inflexible or rigid  -QI intervention has perceived negative unintended or unanticipated consequences e.g., creates more administrative burden on already overstretched health staff  -Intervention does not allow implementers (who see it as alien or imposed upon them) to make or suggest adaptations  -intervention package does not envisage nor address other contextual and systems barriers to its successful implementation (focus on short term technical fixes and does not address or consider structural bottlenecks)  -QI intervention does not build on existing initiatives | **Africa (Low-income):** Hounsou et al (2022) - Benin; Coulibaly et al (2020) - Mali; Gage et al (2022) - Zimbabwe; Stover et al (2014) - Ethiopia; Quaife et al (2021) - Ethiopia; Ayele et al (2019) - Ethiopia; Tiruneh et al (2020) - Ethiopia; Tibeihaho et al (2021) - Uganda; Kim et al (2019) - Uganda; Hutchinson et al. (2021) - Uganda; Werdenberg et al (2018) - Rwanda; Umunyana et al (2020) - Rwanda  **Africa (Lower middle-income):** Giessler et al (2020) - Kenya; Eboreime et al (2018) - Nigeria; Eboreime et al (2019) - Nigeria; Olaniran et al (2022) – Nigeria; Tancred et al (2016) - Tanzania; Jaribu et al (2017) - Tanzania; Tancred et al (2018) - Tanzania; Pallangyo et al (2018) - Tanzania; Baker et al (2018) - Tanzania;  **Africa (Upper middle-income)**: Basenero et al (2022 - Namibia; Yapa et al (2022) - South Africa; Mantell et al (2022) - South Africa; Mutambo et al (2020) - South Africa; Kinney et al (2022) – South Africa; Horwood et al (2023) - South Africa  **Asia (Upper middle-income)**: Lall et al (2020) - India; Schierhout et al (2021) - India; Werner et al (2021) - Tajikistan; Schuele and MacDougall (2022) - Papua New Guinea; Thekkur et al (2022) - Sri Lanka; Limato et al (2019) - Indonesia  **Americas (Low-income):** Demes et al (2021) - Haiti  **Americas (Upper middle-income)**: Pesec et al (2021) - Costa Rica  **Multi-country:** Sukums et al (2015) - Tanzania and Ghana; Kinney et al (2020) - Tanzania, Nigeria, Rwanda, Zimbabwe; Chandani et al 2017) - Rwanda and Malawi; Djellouli et al (2016) - Malawi, Kenya, Burkina Faso and Mozambique |
| Organization and Team implementing QI | *Enablers:*  -managers and team members agree to additional responsibilities  -seniour leaders embrace and support QI  -experienced subject matter experts drive change  -collegiality or team spirit in decision making beginning from the start of QI project  -presence of QI champions in the team  -balance between top-down and bottom-up approaches in decision making  -team enthusiastic and (publicly) committed  -everyone involved with diverse inputs  -a quality culture with shared values, attitudes and behaviour of everybody becomes embedded in the organisation’s fabric e.g., regular data analysis, action and improvement cycles  -organization allocates budget, avails resources for QI  -physicians take lead, build others’ skills  -trained team members report back, share knowledge and skills with colleagues e.g., on Plan-Do-Study-Act cycles and problem-solving  -regular, positive feedback on QI project shared with stakeholders including good internal communication  -positive team experiences from successful legacy QI projects produce domino effect  -adequate team preparation before introduction of QI  -regular on-the-job training in addition to classroom sessions  -accreditation process inspires and supports drive to improve service quality  *Barriers:*  -frozen relationships between managers and frontline implementers  -organization does not own (rejects) new QI initiative  -team members lack knowledge or skills on QI approaches  -lack of clarity on QI stewardship and monitoring arrangements  -The ‘missing middle’ in decentralized settings (unsupportive district-level managers)  -concurrent similar QI programmes in the same organization bring confusion and uncertainty  -team neglects to include support (non-technical) staff  -team leaders do not genuinely involved others in decisions  -weak leadership by government sees QI left to partners/donors  -one-off training for QI team norms  -QI focal persons wearing too many hats | **Africa (Low income):**  Coulibaly et al (2020) - Mali  Nahimana et al (2021) - Rwanda; Umunyana et al (2020) - Rwanda; Stover et al (2014) - Ethiopia  **Africa (Lower middle-income)**: Eboreime et al (2018) – Nigeria; Baker et al (2018) - Tanzania; Pallangyo et al (20180 - Tanzania  **Africa (Upper middle-income):** Kinney et al (2022) – South Africa; Mantell et al (2022) - South Africa; Yapa et al (2022) - South Africa; Horwood et al (2023) - South Africa  **Asia (Upper middle-income):** Schierhout et al (2021) - India; Limato et al (2019) - Indonesia; Schuele and MacDougall (2022) - Papua New Guinea; Werner et al (2021) - Tajikistan  **Americas (Low income)**: Demes et al (2021) - Haiti  **Americas (Upper middle-income):** none  **Multi-country:** Kinney et al (2020) - Rwanda, Tanzania, Zimbabwe, Nigeria; Chandani et al (2017) – Rwanda and Malawi |
| Health Systems Support and Capacity | *Enablers:*  -available staff with aligned job descriptions and incentives  -adequate, well designed physical space and infrastructure  -facilitative and supportive supervision  -regular follow up and mentorship  -silos and lack of integration  -provision of adequate supplies and commodities to deliver services  -strong patient referral  -participatory and data-driven QI activities  -data and reporting tools are revised to ensure one harmonized system of reports  *Barriers:*  -stockouts of drugs and supplies  - inadequate patient referral systems  -unpredictable follow up and punitive or unfocused supervision  -frequent staff leave of absence  -high staff turnover at health facility  -low numbers of health workers with high work loads  -poorly designed or inadequate space and infrastructure  -lack of equipment (ICT/data and medical devices)  -insufficient engagement of district level  -inadequate patient records system at the health facility level constrains service delivery | **Africa (Low-income):** Manzi et al (2014) - Rwanda; Tayebwa et al (2020) - Rwanda; Nahimana et al (2021) - Rwanda; Umunyana et al (2020) - Rwanda; Werdenberg et al (2018) - Rwanda; Bradley et al (2012), Ethiopia; Stover et al (2014), Ethiopia; Ayele et al (2019), Ethiopia; Coulibali et al (2020) - Mali; Hounsou et al (2022) - Benin  **Africa (Lower middle-income):** Eboreime et al (2018) - Nigeria; Olaniran et al (2022) – Nigeria; Baker et al (2018) - Tanzania; Pallangyo et al (2018) - Tanzania  **Africa (Upper middle-income):** Kinney et al (2022) – South Africa; Yapa et al (2022) - South Africa; Horwood et al (2023) - South Africa; Mantell et al (2022) - South Africa; Basenero et al (2022 - Namibia  **Asia (Upper middle-income):** Thekkur et al (2022) - Sri Lanka; Schierhout et al (2021) - India; Werner et al (2021) - Tajikistan; Limato et al (2019) - Indonesia  **Americas:** none  **Multi-country:** Chandani et al (2017) - Rwanda and Malawi; Sukums et al (2015) - Tanzania and Ghana; Djellouli et al (2016) - Malawi, Kenya, Burkina Faso and Mozambique; Kinney et al (2020) - Rwanda, Tanzania, Zimbabwe and Nigeria |
| External environment and structural factors | *Enablers:*  -needed policies, plans, budgets and guidelines in place and conducive  -conducive financing and technical policies and guidelines  -high political visibility for QI intervention  -social norms encourage positive collaboration, problem solving and success  -strong political commitment for change  *Barriers:*  -difficult access to/for communities with poor road networks  -conflicts and insecurity, drought and famine  -bad political and socio-economic policies  -international and donor-led priority-setting  -PHC not prioritised - more focus on secondary and tertiary care by government and international agencies  -financial access barriers and poverty  -donor-driven priority setting  -larger health systems configuration e.g., employment conditions and administrative set up  -poor roads, energy & telecommunications infrastructure  - poor weather conditions  -disruptive onset of COVID-19 pandemic  -weak regulation and integration of private PHC service providers in health system  -weak collaboration and coordination between central and peripheral (local) government structures | **Africa (Low-income):** Lokossou et al (2019) - Benin; Coulibaly et al (2020) - Mali; Bradley et al (2012) - Ethiopia; Nahimana et al (2021) - Rwanda; Werdenberg et al (2018) - Rwanda  **Africa (Lower middle-income):** Olaniran et al (2022) - Nigeria  **Africa (Upper middle-income):** Yapa et al (2022) - South Africa; Horwood et al (2023) - South Africa; Mantell et al (2022) - South Africa; Mutambo et al (2020) - South Africa; Kinney et al (2022) - South Africa  **Asia (Upper middle-income):** Werner et al (2021) - Tajikistan; Thekkur et al (2022) - Sri Lanka  **Americas:** none  **Multi-country:** Djellouli et al (2016) - Kenya, Malawi, Mozambique, Burkina Faso; Sukums et al (2015) - Tanzania and Ghana; Kinney et al (2020) - Rwanda, Tanzania, Zimbabwe, Nigeria |
| Execution of QI Intervention | *Enablers:*  -Implementers work collaboratively with community resource persons and civil society, draw upon local knowledge to tailor communication to clients and to effectively engage with communities  -champions are identified across all levels of the organization and system and take lead on modelling new roles in PHC while emphasizing collaborative working  -adequate numbers of implementers receive ongoing knowledge and practice updates from knowledgeable mentors and supervisors, and supervision/mentorship sessions embrace reflexivity and reflective practice.  -unconstrained communication makes use of multiple channels, provides avenue for (real-time) feedback and information sharing across all levels and types of QI stakeholders and facilitates decision-making  -including reminders in home-based records for patients where applicable  -re-designing clinic workflow, as needed, in a patient-centered manner  -stocks of key commodities are tracked and reported regularly  -results-oriented work plans are developed and executed participatorily  -QI implementation includes enhancements in documentation of care processes  -intervention is executed in incremental doses where subsequent sessions build on earlier ones in a responsive manner  -there is verification (monitoring) of whether QI activities are implemented in line with plans using data from PHC facilities  -influencers and blockers are identified and engaged during QI implementation  -QI training sessions are offered repeatedly to reach most implementers  *Barriers:*  -QI implementation does not consider availability of staff and competing tasks, leading to some health workers missing meetings and training sessions  -focus of intervention remains limited throughout implementation period, and not all planned aspects get rolled out. Late roll out of only a few aspects.  -clients keep off PHC facilities due to past negative experiences when seeking care  -implementation plans considered over-ambitious and unrealistic  -limited training and supervision of health service providers create gaps in implementation  -community clients stay away due to low or non-involvement of local leaders and administrators exposing only a few clients to the QI intervention that targets them  -implementers withhold feedback from other stakeholders including communities contributing to mistrust, misperceptions, and constrained relationships  -lack of support supervision during QI implementation  -objectives of QI sessions are not discussed or shared widely  -limited risk communication and communities remain unaware of the need to shift behaviours and practices to healthier options promoted by QI intervention  -implementers do not keep track of the availability of drugs and other stocks  -implementation is skewed away from agreed plans to meet donor demands  -health workers do not practice new skills gained from QI for extended periods leading to decay of knowledge and skills  -users (in case of technology) experience delays when stuck and need support | **Africa (Low-income):** Coulibaly et al (2020) - Mali; Hounsou et al (2022) - Benin; Stover et al (2014) - Ethiopia; Bradley et al (2012) - Ethiopia; Ayele et al (2019) - Ethiopia; Quaife et al (2021) - Ethiopia; Manzi et al (2014) - Rwanda; Werdenberg et al (2018) - Rwanda; Nahimana et al (2021) - Rwanda; Umunyana et al (2020) - Rwanda; Tayebwa et al (2020) - Rwanda; Hutchinson et al (2021) - Uganda  **Africa (Lower middle-income):** Eboreime et al (2018) - Nigeria; Olaniran et al (2022) – Nigeria; Jaribu et al (2016) - Tanzania; Pallangyo et al (2018) - Tanzania; Tancred et al (2018) - Tanzania; Baker et al (2018) - Tanzania  **Africa (Upper middle-income):** Basenero et al (2022 - Namibia; Yapa et al (2022) - South Africa; Mantell et al (2022) - South Africa; Mutambo et al (2020) - South Africa; Horwood et al (2023) - South Africa; Kinney et al (2022) - South Africa  **Asia (Upper middle-income):** Thekkur et al (2022) - Sri Lanka; Limato et al (2019) - Indonesia; Schierhout et al (2021) - India; Werner et al (2021) - Tajikistan  **Americas:** none  **Multi-country:** Djellouli et al (2016) - Kenya, Malawi, Mozambique, Burkina Faso; Kinney et al (2020) - Tanzania, Nigeria, Rwanda, Zimbabwe; Chandani et al (2017) - Rwanda and Malawi; Sukums et al (2015) - Tanzania and Ghana |
