## Supplemental Table 6. Topics covered in QI research for "Barriers to and enablers of quality improvement in primary health care in low- and middle-income countries: a systematic review"

**S6 Table. Quality improvement research topics and approaches in LMICs**

| **Research area** | **Frameworks and models** | **Methods** | **Studies** |
| --- | --- | --- | --- |
| Continuous quality improvement/ quality improvement collaborative | Force field analysis, derived from Kurt Lewin’s force field theory; PDSA cycles. | Semi-structured interviews (SSIs) and document reviews- deductive thematic analysis; FGDs, KIIs, document reviews and health systems performance data- Grounded theory; interviews and FGDs- thematic analysis. | Gage et al (2022) - Zimbabwe; Tibeihaho et al (2021) - Uganda; Limato et al (2019) - Indonesia; Schuele and MacDougall (2022) - Papua New Guinea |
| Digital health interventions | COM-B Theory of Change model; Implementation research framework; RE-AIM framework. | SSIs and group discussions- inductive and deductive coding; Questionnaires, Administrative data, IDIs and FGDs- descriptive statistics and deductive thematic analysis; Interviews, FGDs- cluster-level framework matrix analysis; Questionnaires, IDIs, supervision checklists, field notes of observations and discussions, and project diaries- Pearson’s Chi-squared test, Fisher’s exact test and thematic analysis. | Sukums et al (2015) - Tanzania and Ghana; Horwood et al (2023) - South Africa; Schierhout et al (2021) - India; Demes et al (2021) - Haiti |
| HIV/AIDS | Root cause analysis (RCA); Normalization process theory. | Document reviews, facility audits, patient satisfaction surveys, focus group interviews, field notes and a reflection diary- inductive analysis; field notes, document reviews and SSIs- framework analysis; IDIs- framework analysis; IDIs- thematic data analysis. | Basenero et al (2022) - Namibia; Mutambo et al (2020) - South Africa; Visser et al (2018) - South Africa; Yapa et al (2022) - South Africa |
| Malaria | PDSA cycles | Ethnographic observations, informal discussions, IDIs, and FGDs- interpretive analysis inductively and deductively. | Hutchinson et al (2021) - Uganda |
| Maternal newborn health | IHI’s Collaborative Model for Achieving Breakthrough Improvement; Barth’s transactional model of culture; Gidden’s Structuration Theory; Promoting Action on Research Implementation in Health Services (PARIHS); IHI’s Breakthrough Series and Model for Improvement; PDSA cycles; Fishbone and Pareto charts. | IDIs- TCA; FGDs- inductive coding; IDIs- Thematic analysis; observation and interviews- descriptive open coding; IDIs and FGDs- content analysis; Interviews- TCA; routinely kept records and birth narratives- deductive thematic analysis; IDIs- content analysis; Questionnaire and IDIs- bivariate and descriptive statistics, constant comparative; birth narratives, IDIs and FGDs- deductive coding; observations, IDIs- deductive thematic analysis; HMIS data, patient charts, CHW records, facility surveys, FGDs and interviews- Wilcoxon signed rank test and inductive coding; review of documents, KIIs, observations- thematic analysis; FGDs, IDIs- inductive-deductive coding; surveys, routine monitoring tools, discussions, FGDs- Chi-squared tests, t-tests, thematic (inductive-deductive) coding; FGDs- thematic analysis. | Tiruneh et al (2020) - Ethiopia; Patterson et al (2021) - Malawi; Jaribu et al (2016) - Tanzania; Tancred et al (017) - Tanzania; Tancred et al (2018) - Tanzania; Stover et al (2014) - Ethiopia; Nahimana et al (2021) - Rwanda; Kim et al (2019) - Uganda; Werdenberg et al (2018) - Rwanda; Pallangyo et al (2018) - Tanzania; Bogren et al (2021) - DRC; Manzi et al (2014) - Rwanda  Giessler et al (2020) - Kenya; Olaniran et al (2021) - Nigeria; Vail et al (2018a) - India; Vail et al (2018b) - India; Djellouli et al (2016) - Burkina Faso, Kenya, Malawi, Mozambique; |
| Maternal perinatal death surveillance and response | MPDSR continuous action cycles; 6-step MPDSR audit cycle; Carl May’s extended normalization process theory. | Document reviews, Semi-structured questionnaire and observations- TCA; semi-structured questionnaire- TCA; semi-structured questionnaire, desk review, KII and observations; individual and group interviews and non-participant observations- thematic analysis; document reviews, group discussions, administrative data- descriptive analysis and inductive content analysis. | Ayele et al (2019) - Ethiopia; Hounsou et al (2022) - Benin; Kinney et al (2022) - South Africa; Tayebwa et al (2020) - Rwanda; Kinney et al (2020) - Rwanda, Tanzania, Zimbabwe, Nigeria |
| Non-communicable diseases | Consolidated Framework for Implementation Research (CFIR); the Model for Understanding Success in Quality (MUSIQ); Tailored Implementation for Chronic Diseases (TICD) network. | IDIs- inductive and deductive coding; IDIs, observations and field notes- framework analysis; SSIs- content analysis. | Wakida et al (2019) - Uganda; Odusola et al (2016) - Nigeria; Basenero et al (2022) - Namibia; Lall et al (2020) - India; Schierhout et al (2021) - India |
| Primary health care systems strengthening | Breakthrough series for collaborative QI; Diagnose-Intervene-Verify-Adjust (DIVA) derived from PDSA cycles; Positive deviance; CFIR; Battacharya et al’s systems approach; Data to Improvement Pathway; the Adaptive Management Framework. | IDIs- inductive and deductive coding using framework analysis; SSIs-thematic analysis; Interviews- Qualitative content analysis; KII-IDIs and HMIS data- inductive and deductive coding; interviews, observations, administrative data- case study analysis; KIIs, IDIs, field observations, surveys, time motion studies, online surveys, and on site assessments- TCA; Self-assessment and document reviews, IDIs- frequencies and percentages, thematic analysis; KIIs- QCA; SSIs and participant observations- deductive–inductive thematic analysis; HMIS and IDIs- linear and non-linear regression, constant comparative; IDIs-exploratory factor analysis after inductive coding; document reviews, KIIs, IDIs- Framework analysis. | Baker et al (2018) - Tanzania; Bradley et al (2012) - Ethiopia; Quaife et al (2021) - Ethiopia; Umunyana et al (2020) - Rwanda; Lokossou et al (2019) - Benin; Coulibaly et al (2020) - Mali; Chandani et al (2017) - Rwanda and Malawi; Eboreime et al (2018) - Nigeria; Eboreime et al (2019) - Nigeria;; Mantell et al (2022) - South Africa; Pesec et al (2021) - Costa Rica; Limato et al (2019) - Indonesia; Thekkur et al (2022) - Sri Lanka; Werner et al (2021) - Tajikistan |
