## Supplemental Figure 4. ProQuest search for "Barriers to and enablers of quality improvement in primary health care in low- and middle-income countries: a systematic review"

| | | | |
|---|---|---|---|
| | tadjikistan OR tadzhikistan OR tadzhik OR tanzania OR tanganyika OR thailand OR siam OR timor leste OR east timor OR togo OR togolese republic OR tonga OR "trinidad and tobago" OR trinidad OR tobago OR tunisia OR turkey OR turkmenistan OR turkmen OR uganda OR ukraine OR uruguay OR uzbekistan OR uzbek OR vanuatu OR new hebrides OR venezuela OR vietnam OR viet nam OR middle east OR west bank OR gaza OR palestine OR yemen OR yugoslavia OR zambia OR zimbabwe OR northern rhodesia OR global south OR africa south of the sahara OR sub-saharan africa OR subsaharan africa OR africa, central OR central africa OR africa, northern OR north africa OR northern africa OR magreb OR maghrib OR sahara OR africa, southern OR southern africa OR africa, eastern OR east africa OR eastern africa OR africa, western OR west africa OR western africa OR west indies OR indian ocean islands OR caribbean OR central america OR latin america OR "south and central america" OR south america OR asia, central OR central asia OR asia, northern OR north asia OR northern asia OR asia, southeastern OR southeastern asia OR south eastern asia OR southeast asia OR south east asia OR asia, western OR western asia OR europe, eastern OR east europe OR eastern europe) AND ("quality improvement" N10 health) AND pd(20000101-20230204) AND pd(>20001231) | | |
| S7 | summary(Barrier* OR limitation* OR constraint* OR enabler* OR promoter* OR facilitator* OR motiv*) AND summary(afghanistan OR albania OR algeria OR american samoa OR angola OR "antigua and barbuda" OR antigua OR barbuda OR argentina OR armenia OR armenian OR aruba OR azerbaijan OR bahrain OR bangladesh OR barbados OR republic of belarus OR belarus OR byelarus OR belorussia OR byelorussian OR belize OR british honduras OR benin OR dahomey OR bhutan OR bolivia OR "bosnia | Acta Sanctorum, American Periodicals, Art, Design & Architecture Collection, Art & Architecture Archive, Avery Index to Architectural Periodicals, British Periodicals, C19: The Nineteenth Century Index, Colonial State Papers, Coronavirus Research Database, Country Life Archive, Digital National Security Archive, Dissertations & Theses @ Lancaster University, Documents on British Policy Overseas, Early Modern Books, Ebook Central, Education Magazine Archive, Entertainment Industry Magazine Archive, LGBT Magazine Archive, Linguistics and Language Behavior Abstracts (LLBA), News, Policy & Politics Magazine Archive (feat. Newsweek), Patrologia Latina, Periodicals Archive Online, ProQuest Dissertations & Theses Global, ProQuest Historical Newspapers: Atlanta Daily World, ProQuest Historical Newspapers: Chicago Defender, | 53 |

| | | |
|---|---|---|
| | and herzegovina" OR bosnia OR herzegovina OR botswana OR bechuanaland OR brazil OR brasil OR bulgaria OR burkina faso OR burkina fasso OR upper volta OR burundi OR urundi OR cabo verde OR cape verde OR cambodia OR kampuchea OR khmer republic OR cameroon OR cameron OR cameroun OR central african republic OR ubangi shari OR chad OR chile OR china OR colombia OR comoros OR comoro islands OR iles comores OR mayotte OR democratic republic of the congo OR democratic republic congo OR congo OR zaire OR costa rica OR "cote d'ivoire" OR "cote d’ ivoire" OR cote divoire OR cote d ivoire OR ivory coast OR croatia OR cuba OR cyprus OR czech republic OR czechoslovakia OR djibouti OR french somaliland OR dominica OR dominican republic OR ecuador OR egypt OR united arab republic OR el salvador OR equatorial guinea OR spanish guinea OR eritrea OR estonia OR eswatini OR swaziland OR ethiopia OR fiji OR gabon OR gabonese republic OR gambia OR "georgia (republic)" OR georgian OR ghana OR gold coast OR gibraltar OR greece OR grenada OR guam OR guatemala OR guinea OR guinea bissau OR guyana OR british guiana OR haiti OR hispaniola OR honduras OR hungary OR india OR indonesia OR timor OR iran OR iraq OR isle of man OR jamaica OR jordan OR kazakhstan OR kazakh OR kenya OR "democratic people's republic of korea" OR republic of korea OR north korea OR south korea OR korea OR kosovo OR kyrgyzstan OR kirghizia OR kirgizstan OR kyrgyz republic OR kirghiz OR laos OR lao pdr OR "lao people's democratic republic" OR latvia OR lebanon OR lebanese republic OR lesotho OR basutoland OR liberia OR libya OR libyan arab jamahiriya OR lithuania OR macau OR macao OR republic of north macedonia OR macedonia OR madagascar OR malagasy republic OR malawi OR nyasaland OR malaysia OR malay federation OR malaya federation OR maldives OR indian ocean islands OR indian ocean OR mali OR malta OR micronesia OR federated states of micronesia OR | ProQuest Historical Newspapers: Chicago Tribune, ProQuest Historical Newspapers: Chinese Newspapers Collection, ProQuest Historical Newspapers: Communist Historical Newspaper Collection, ProQuest Historical Newspapers: Leftist Newspapers and Periodicals, ProQuest Historical Newspapers: Le Monde, ProQuest Historical Newspapers: Los Angeles Sentinel, ProQuest Historical Newspapers: Michigan Chronicle, ProQuest Historical Newspapers: New York Amsterdam News, ProQuest Historical Newspapers: New York Tribune / Herald Tribune, ProQuest Historical Newspapers: Norfolk Journal and Guide, ProQuest Historical Newspapers: Philadelphia Tribune, ProQuest Historical Newspapers: Pittsburgh Courier, ProQuest Historical Newspapers: South China Morning Post, ProQuest Historical Newspapers: The Baltimore Afro-American, ProQuest Historical Newspapers: The Globe and Mail, ProQuest Historical Newspapers: The Guardian and The Observer, ProQuest Historical Newspapers: The Jerusalem Post, ProQuest Historical Newspapers: The Korea Times, ProQuest Historical Newspapers: The New York Times with Index, ProQuest Historical Newspapers: The Times of India, ProQuest Historical Newspapers: The Washington Post, ProQuest One Business, ProQuest One Literature, Publicly Available Content Database, Religious Magazine Archive, The Artforum Archive, The Harper's Bazaar Archive, The Rolling Stone Archive, The Vogue Archive, The Women's Wear Daily Archive, Trench Journals and Unit Magazines of the First World War, Women's Magazine Archive, Youth and Popular Culture Magazine Archive |

Database copyright © 2023 ProQuest LLC. All rights reserved.

Terms and Conditions Contact ProQuest
